# A hidden Markov model reliably characterizes ketamine-induced spectral dynamics in macaque LFP and human EEG

**DOI:** 10.1101/2020.11.12.20221366

**Authors:** Indie C. Garwood, Sourish Chakravarty, Jacob Donoghue, Pegah Kahali, Shubham Chamadia, Oluwaseun Akeju, Earl K. Miller, Emery N. Brown

## Abstract

Ketamine is an NMDA receptor antagonist commonly used to maintain general anesthesia. At anesthetic doses, ketamine causes bursts of 30-50 Hz oscillations alternating with 0.1 to 10 Hz oscillations. These dynamics are readily observed in local field potentials (LFPs) of non-human primates (NHPs) and electroencephalogram (EEG) recordings from human subjects. However, a detailed statistical analysis of these dynamics has not been reported. We characterize ketamine’s neural dynamics using a hidden Markov model (HMM). The HMM observations are sequences of spectral power in 10 Hz frequency bands between 0 to 50 Hz, where power is averaged within each band and scaled between 0 and 1. We model the observations as realizations of multivariate beta probability distributions that depend on a discrete-valued latent state process whose state transitions obey Markov dynamics. Using an expectation-maximization algorithm, we fit this beta-HMM to LFP recordings from 2 NHPs, and separately, to EEG recordings from 9 human subjects who received anesthetic doses of ketamine. Together, the estimated beta-HMM parameters and optimal state trajectory revealed an alternating pattern of states characterized primarily by gamma burst and slow oscillation activity, as well as intermediate states in between. The mean duration of the gamma burst state was 2.5s([1.9,3.4]s) and 1.2s([0.9,1.5]s) for the two NHPs, and 2.7s([1.9,3.8]s) for the human subjects. The mean duration of the slow oscillation state was 1.6s([1.1,2.5]s) and 0.7s([0.6,0.9]s) for the two NHPs, and 2.8s([1.9,4.3]s) for the human subjects. Our beta-HMM framework provides a useful tool for experimental data analysis. Our characterizations of the gamma-burst process offer detailed, quantitative constraints that can inform the development of rhythm-generating neuronal circuit models that give mechanistic insights into this phenomenon and how ketamine produces altered states of arousal.

## 1 Introduction

Ketamine is a phenylcyclidine derivative and one of the most commonly used anesthetics in clinical use worldwide [1, 2, 3, 4]. It is classified as a WHO Essential Medicine [5], and is often the only anesthetic available in clinics of developing countries [6]. At low doses, ketamine is known to create a state of “dissociative anesthesia” characterized by altered sensory perception and analgesia [1, 2]. At high doses it also leads to loss of consciousness, and therefore it is often used as an anesthetic agent during surgery in humans and animals [3, 7, 8, 9]. Under anesthetic dosages, ketamine produces distinct oscillatory signatures in the electroencephalogram (EEG) of healthy volunteers and patients [10, 11, 12]. In a recent retrospective study by Akeju et al. [11], the authors found that the frontal EEG from patients who received ketamine for the induction of general anesthesia showed distinct alternating periods of intermittent high power activity in the 27 − 40 Hz and 0.1 − 4 Hz frequency bands. Spontaneous gamma oscillations (i.e. 30 − 50 Hz) have also been demonstrated under ketamine-induced general anesthesia in mice [13], non-human primates (NHPs) [14, 15, 16], and sheep [17]. Recent neuroscience research in NHPs under high-dose ketamine, revealed prominent gamma oscillations modulated by slow wave (0.3 Hz) activity [16], similar to the alternating bursts of activity in gamma and slow-delta bands observed in humans [11]. Objective characterization of the intermittent band-limited spectral dynamics induced by ketamine would be useful both for monitoring patients during ketamine-induced unconsciousness in clinical settings, as well as for aiding computational and experimental neuroscience research aimed at building a mechanistic understanding of the phenomena. Therefore, the availability of an efficient analytic tool, which can objectively detect and characterize ketamine-induced transient neural dynamics, can catalyze both clinical and neuroscience innovations.

Ketamine’s primary pharmacologic effect is N-Methyl-d-aspartate (NMDA) receptor antagonism. One putative mechanism of ketamine induced cortical gamma oscillations is that ketamine preferentially blocks NMDA receptors on fast spiking inhibitory gamma aminobutyric acid (GABA) interneurons. The blocking of the NMDA receptors on the interneurons results in reduced interneuron activity, and a reduction of GABA release at the synapse between interneurons and excitatory pyramidal neurons. The reduction of GABA causes disinhibition of the pyramidal neurons, resulting in increased pyramidal neuron activity that manifests as cortical gamma oscillations [18, 19]. This mechanism, however, does not explain the presence of periodic bursts of gamma activity. A recent modeling study suggests a cyclic mechanism where in each cycle, the pyramidal neurons that are disinhibited at the beginning of the cycle provide sufficient input to the interneurons for the latter to overcome the ketamine inhibition and release GABA, which then results in temporary inhibition of the pyramidal neurons towards the end of the cycle [20]. A statistical characterization of the ketamine-induced alternating gamma burst and slow-oscillation activity in neural data can provide informative quantitative constraints for mechanistic neuronal circuit models of ketamine-induced spectral dynamics. Such mechanistic models can further aid the understanding of how ketamine causes altered states of arousal. Examples of how detailed statistical analyses of neural data has guided neurophysiological modeling studies can be found in existing works that studied propofol-induced unconsciousness [21, 22, 23, 24].

State-space models provide a versatile statistical framework for analyzing the dynamics of complex neural time-series data [25]. These models have been widely used in neuroscience, from decoding hippocampal place cells [26], to tracking movement intention in the motor cortex [27, 28, 29], to assessing the level of unconsciousness in medically-induced coma [30]. A particular class of state-space models, known as Hidden Markov Models (HMMs), have been used to describe underlying dynamical processes driven by stochastic switching between discrete states, where each state is associated with a distinct observation probability distribution [31, 32, 33, 34, 35, 36]. HMMs have been used to objectively decode neurophysiological states, including detecting seizure events [37], classifying sleep stages [38, 39, 40, 41], decoding speech [42, 43], and decoding movement intention [44, 45]. HMMs have also been used to provide detailed statistical descriptions of neural phenomena [46, 47, 48]. Since ketamine-induced cortical activity is characterized by distinct switching patterns in oscillatory dynamics, an HMM is a plausible statistical model for both estimating neurophysiological states and characterizing their statistical properties. Our preliminary analysis using an HMM to characterize ketamine-induced spectral dynamics in human EEG and in local field potential (LFP) from NHPs yielded promising results [49]. Along similar lines, the recent work by Li and Mashour [12], using a particular class of HMMs [50] to characterize multichannel scalp EEG from healthy volunteers, further supports the utility of HMMs to analyze ketamine spectra. By inferring an HMM from neural activity spectra, we can quantitatively characterize the discontinuous spectral dynamics in terms of the parameters of the best-fitting observation and state transition models, as well as the state trajectory corresponding to the entire data sequence.

In this work, we develop an HMM-based analysis framework to characterize ketamine-induced spectral dynamics recorded in neurophysiological data from 2 NHPs and 9 human subjects in the operating room (see schema in Fig. 1). The observations that go into our HMM analysis are sequences of spectral power in 10 Hz frequency bands between 0 and 50 Hz. The power is averaged within each band and scaled between 0 and 1 to facilitate comparison among data sets that vary in power amplitude but not in the spectral profiles they represent. To describe this multivariate observation sequence, we assume that the spectral dynamics are governed by a discrete-valued latent state process, where each state corresponds to a distinct spectral profile. We model these spectral profiles as realizations of state-specific and frequency-band specific beta distributions. From the estimated HMM, we are able to objectively characterize the dynamic evolution of neurophysiological states induced by ketamine. By applying this tool to analyze neural data from both NHPs and humans, we obtain precise estimates of time-scales associated with neurophysiological phenomena observed in both species (e.g., alternating gamma-slow activity).

**Figure 1:**
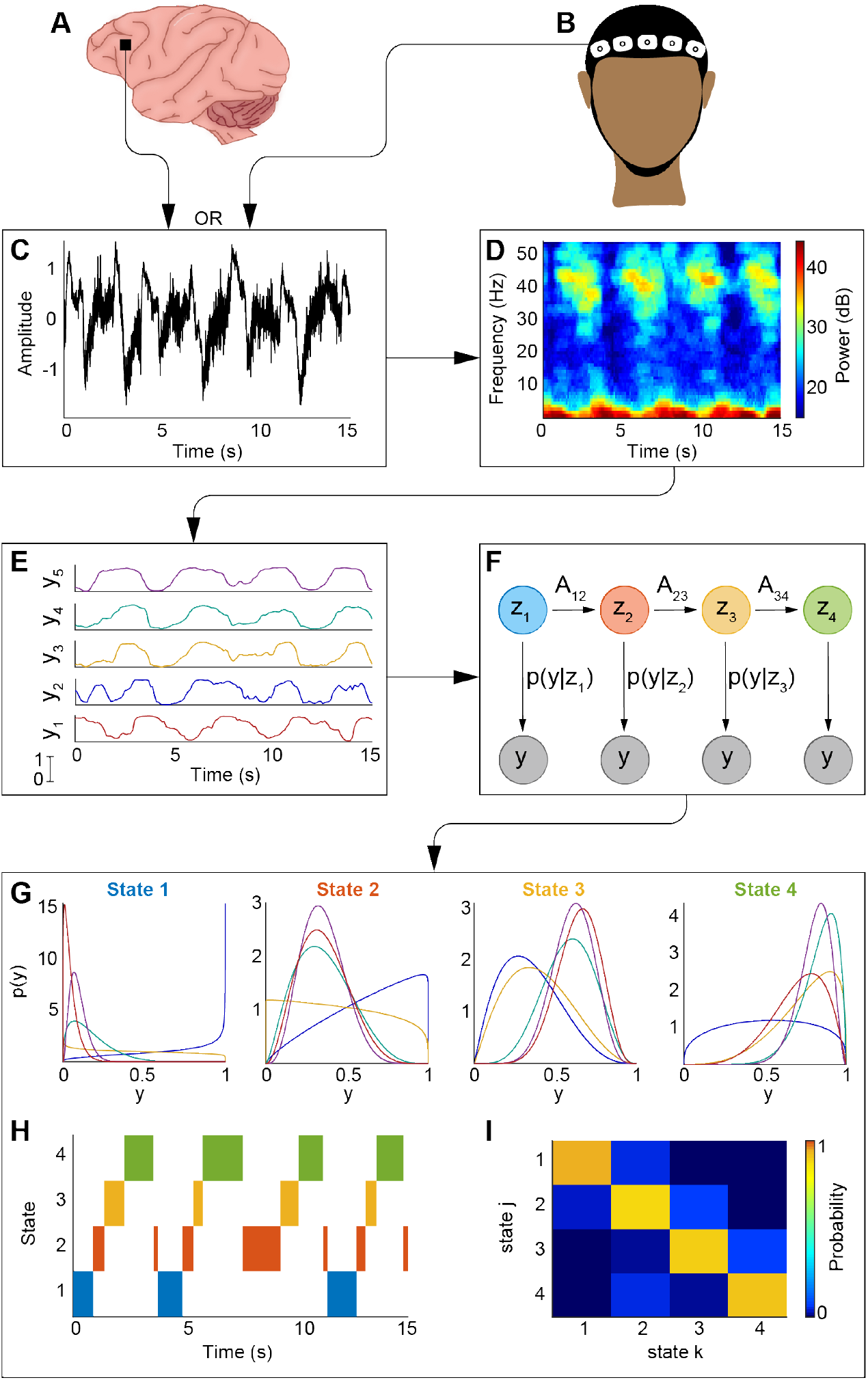
Schematic of the beta-HMM algorithm. Neural recordings from primate LFP (A) or human EEG (B) serve as inputs to the algorithm. Electrical recordings (C) are converted to their time-frequency representation via multitaper spectral analysis (D). The spectral observations are split into 10 Hz frequency bands and scaled between 0 and 1 (E). These observations are then fit to the beta-HMM (F) via the EM algorithm, and the state trajectory is estimated from the Viterbi algorithm. Outputs of the algorithm are the beta observation distributions for each state (G), estimated state trajectory (H), and a transition matrix (I) describing the state switching dynamics.

The paper is organized as follows. In the following section 2 we present our beta-HMM formulation, corresponding estimation framework, and useful statistics based on the estimated beta-HMM. In section 3 we describe the electrophysiology datasets to which we apply our analysis framework. We present the results from our beta-HMM analyses in section 4. Finally, in section 5 we summarize the core contribution of our work, highlight the connections between our work and that of others, and suggest next steps.

## 2 A hidden Markov model for beta distributed observations

### 2.1 Reduced-order observations derived from multitaper spectrogram

We consider a sequence of scalar-valued single-channel voltage activity representing either LFP or EEG sampled at frequency *F*_*s*_ (in Hz). To quantify the spectral dynamics in the signal, we estimate time-varying spectra across a sequence of overlapping time-windows, each of duration Δ_*MT*_ = 1s, with 90% overlap between consecutive windows. The power spectral density (in dB) at the *n*-th time-window and corresponding to frequency *ω*_*i*_ (in Hz) is denoted by *S*_*n*_(*ω*_*i*_). We use the multitaper spectral estimation approach [52, 53, 54] (time-halfbandwidth product = 2, and number of tapers = 3) to calculate the sequence 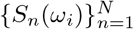 on a high dimensional set of distinct frequencies {*ω*_*i*_ : 0 *< ω*_*i*_ *< F*_*s*_*/*2}. We reduce the complexity of our subsequent analyses by summarizing the high-dimensional information *S*_*n*_({*ω*_*i*_}) by a low-dimensional vector 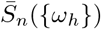. This low-dimensional vector characterizes the instantaneous average power in *H* (= 5) distinct 10 Hz frequency bands between 0-50 Hz, such that

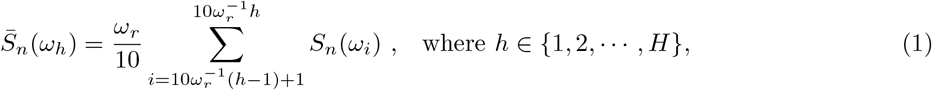

and *ω*_*r*_ denotes the frequency resolution of the multitaper spectral estimates. The choice of the frequency bands was based on our initial observation that in both NHP LFP and human EEG, significant dynamic activity across 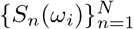 occurred in 0 − 50 Hz during ketamine induction [49]. This range encompasses the frequencies considered in previous studies of ketamine-induced changes in EEG spectra in healthy human subjects and patients [11, 12]. The sequence 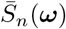 where, 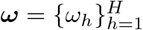, is input to the beta-HMM analysis ^1^.

### 2.2 Observation model

To facilitate the comparison of spectral observations across different data sets whose spectral power may vary in amplitude but represent comparable spectral profiles, we scale 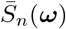 to new vector ***y***_*n*_ ∈ [0, 1]^*H*^ whose *h*-th element is calculated as,

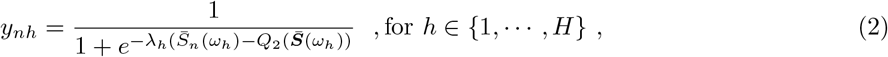

where, 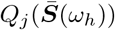 denotes the function that operates on a real-valued sequence 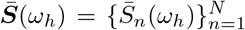 to yield the *j*-th quartile. The user-defined parameter *λ*_*h*_ controls the mutual separation of points in the *y*_*nh*_-space relative to their corresponding map in the 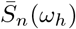-space. We set this value to be 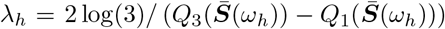. The intuition behind this choice is that it results in scaling of the data between the 1st and 3rd quartiles approximately linearly between [0.25, 0.75]. Data below the 1st quartile and beyond the 3rd quartile are scaled non-linearly, such that very small values are scaled to be close to zero and very large values are scaled to be close to one. Compared to linear 0-1 scaling, the logistic scaling method is less sensitive to outliers which, in linear scaling, can lead to low variance observations and thus reduce the likelihood of detecting distinct spectral profiles. With logistic scaling, the data in 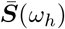 is scaled such that the standard deviations of the observations in each frequency band (***y***_*h*_) are between [0.22, 0.30] for all data sets reported here.

In our model, we assume that for the *n*-th time-window, the observation ***y***_*n*_ is a manifestation of an underlying latent brain state, *z*_*n*_, at the same instant. We also assume that *z*_*n*_ is a random process that transitions among *K* possible discrete states, each with its own characteristic observation probability distribution whose mean corresponds to a distinct spectral profile. We represent a state-specific and frequency band-specific probability density function (pdf) of the observations as a standard beta distribution:

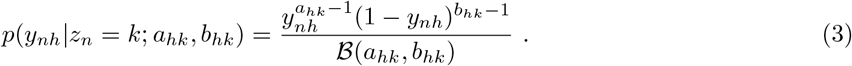

Here, *p*(*y*) denotes the pdf of a continuous-valued random variable *y*. In Eq. (3), *a*_*hk*_ *>* 0 and *b*_*hk*_ *>* 0 are real-valued scalar parameters of the beta distribution that correspond to the scaled power in the *h*-th (*h* ∈ [1, *H*]) frequency band when the state *k* ∈ [1, *K*] is active. The normalization term ℬ (*a*_*hk*_, *b*_*hk*_) denotes the beta function, ℬ (*a*_*hk*_, *b*_*hk*_) = Γ(*a*_*hk*_)Γ(*b*_*hk*_)*/*Γ(*a*_*hk*_ + *b*_*hk*_), where 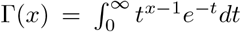 for any scalar real-valued *x >* 0.

Our assumption of statistical independence across the *H* distinct frequency bands conditioned on an underlying state, as implied in Eq. (3), leads to the conditional joint pdf (jpdf) on ***y***_*n*_,

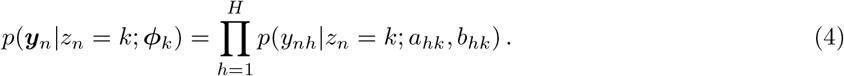

where 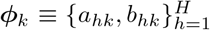 corresponds to the set of 2*H* beta distribution parameters associated with state *k*. The set of beta parameters across all states is denoted as 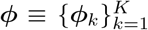. While the beta distribution can be multimodal when both *a*_*hk*_ and *b*_*hk*_ ≤ 1, we require that each state-specific pdf is unimodal. A multimodal pdf associated with a latent state would indicate that the same state is associated with two different spectral profiles, which will confound our intended interpretation of the states. Therefore, to ensure the unimodality in the marginal pdf’s we restrict the domain of the beta distribution parameters to avoid scenarios when both *a*_*hk*_ ≤ 1 and *b*_*hk*_ ≤ 1 for a given *h* and *k*. In summary, we chose to work with the beta distribution because it is a continuous probability distribution defined on the interval [0, 1]. Furthermore, it is part of the exponential family which leads to efficient maximum likelihood estimation (section 2.4). The beta distribution is highly flexible as it allows for unimodal distributions where the modes can lie anywhere in [0, 1].

### 2.3 State transition model

We assume the transition dynamics of the discrete-valued latent state process, *z*_*n*_, to be governed by first-order Markov transitions characterized by a constant transition probability matrix, 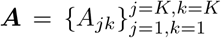, such that 

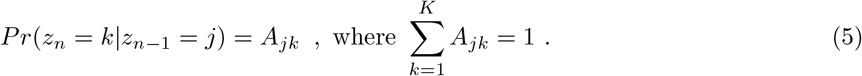

Here, *Pr*(*z*) denotes the probability mass function of a discrete-valued random variable *z*. The initial state probability at the first time-window is characterized by a vector 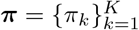 such that

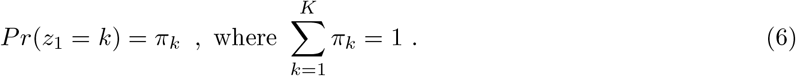

Together, the observation model (Eq. (4)), state transition matrix (Eq. (5)), and initial state probabilities (Eq. (6)) define our state-space model, which we refer to as the *beta-HMM*.

### 2.4 Estimation of states and model parameters

We seek to determine the model parameters of the beta-HMM from one or more single channel LFP or EEG LFP recordings using the maximum likelihood approach via the well-established Expectation-Maximization (EM) algorithm [31, 33, 55, 56]. For a given set of *L* separate recording sessions, {***Y***_1_, …, ***Y***_*L*_}, where the *l*-th session is denoted by 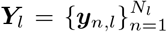, we assume that data can be described by an HMM associated with the parameter set, 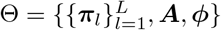. Thus, we assume that the transition matrix, ***A***, and the state-specific beta distribution parameters, 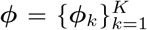, remain constant across all *L* sessions, but the initial state probability ***π***_*l*_ can vary across the sessions. When *L* = 1 the problem reduces to learning a beta-HMM using data from a single recording session as demonstrated later in Sec.4.2 with an LFP recording session from an NHP. In this work (as shown in Sec. 4.3) we also learn subject specific beta-HMMs for 2 NHPs using *L* = 4 separate recording LFP sessions from one NHP, and *L* = 5 from the other. As shown in Sec. 4.4 we also learn a beta-HMM for a typical patient using *L* = 9 independent EEG recording sessions each from a separate human subject. For *L* ≥ 1 mutually independent recording sessions the complete data likelihood for a given Θ can be written as,

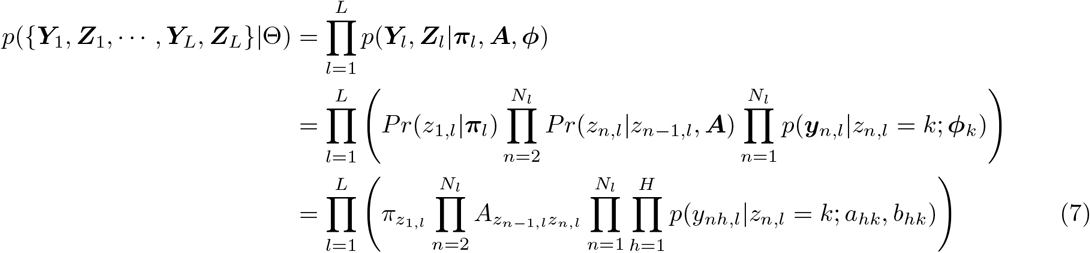

where subscript (·)_,*l*_ denotes the random processes corresponding to the data sequence ***Y***_*l*_. In the above expression, 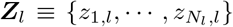 denotes the sequence of latent states corresponding to ***Y***_*l*_. In the ensuing discussion, we shall refer to the expression, log(*p*({***Y***_1_, ***Z***_1_, …, ***Y***_*L*_, ***Z***_*L*_}|Θ)), as the complete data log-likelihood. Note that by marginalizing over all possible {***Z***_1_, …, ***Z***_*L*_}, we are able to obtain the likelihood of {***Y***_1_, …, ***Y***_*L*_} given the model, *p*({***Y***_1_, …, ***Y***_*L*_}|Θ).

The structure of the likelihood expression (Eq. (7)) allows for the maximum likelihood beta-HMM to be estimated in terms of the parameter set 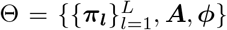 using the EM algorithm [31, 33, 55, 56]. The EM algorithm is commonly used for HMMs (the case of *L* = 1 being the most familiar case [33]) with observation distributions from the exponential family (e.g. the Gaussian distribution) [31, 33, 57, 58]. Briefly, the standard EM algorithm is an iterative optimization scheme where in each EM iteration, we perform an E-step followed by an M-step. In the E-step, following [31, Eq. 13.17], we run *L* independent *Forward-Backward* algorithms ([31, 33]) to estimate the sequences 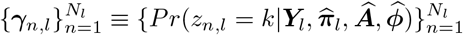 (i.e. the posterior probability of each state at each time-point) and 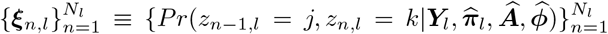 (i.e. the posterior probability of each state transition) given the observations ({***Y***_1_, …, ***Y***_*L*_}) and the last best estimate of the model parameters 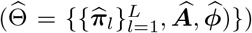 from the previous iteration. Note that the likelihood value, 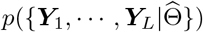, can also be readily calculated following the forward-step of the Forward-Backward algorithm [31, 33].

The M-step of each iteration of the EM algorithm is posed as a maximization (over the space of allowable parameters Θ) of the *expectation* of the complete data log-likelihood function, denoted by 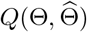 and defined as,

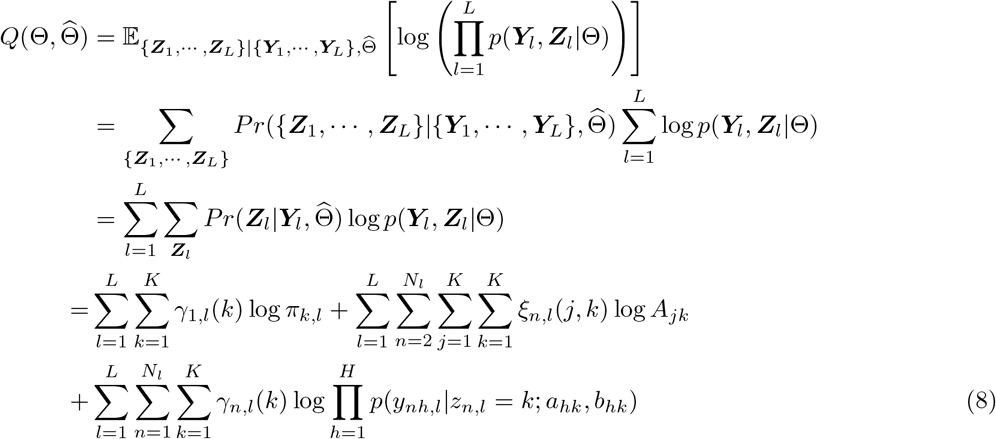

where the last equality follows from Eq. (7). The expectation operation (denoted by 𝔼) indicated in the first equality of Eq. (8) is based on the joint posterior probability distribution on the latent state trajectory estimated using the last best estimate 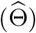 of the HMM. In the M-step, we determine the values for 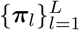, ***A*** and ***ϕ*** that maximize 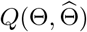 in Eq. (8), which we then use to update 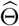. Additional constraints on this maximization problem are given by the equality constraints on ***A*** (Eqs. (5)) and on ***π*** (Eq. (6)), and the following inequality constraints: 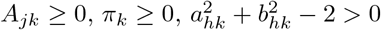 for all *j, k* ∈ [1, *K*] and *h* ∈ [1, *H*]. The last set of inequality constraints on the beta pdf parameters leads to unimodal beta pdf’s.

For an exponential family of observation distributions, the sequence of 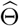’s, resulting from maximizing 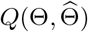 in each EM iteration, leads to a monotonic approach to a maxima of the likelihood function [59, 57, 58, 33]. Therefore, we use the estimated log-likelihood to track EM-algorithm convergence, i.e. we terminate the EM algorithm at an iteration number, *t*, when 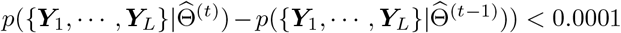. Since the EM algorithm is known to converge to a local maxima [59], we repeat the EM algorithm with multiple initial guesses of Θ and choose the set of model parameters with the highest likelihood. Essential steps relevant to the M-step are presented in the appendix (Sec. A).

Furthermore, using the ML beta-HMM we solve for the optimal state trajectory 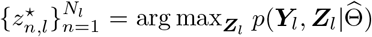 using the *Viterbi* algorithm [31, Sec. 13.2]. This state trajectory represents the unsupervised segmentation of the data sequence, ***Y***_*l*_. The output of the HMM fitted to the multiple observations from multiple sessions are the estimates of the ML parameter set, 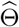, and corresponding sequences of 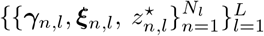. Since the assignment of state label is random, we re-number the states from 1 to *K* in the ascending order of the mean (= *a*_*Hk*_*/*(*a*_*Hk*_ + *b*_*Hk*_)) of the beta distributions corresponding to the *H*-th frequency band.

### 2.5 Statistical analysis using beta-HMM parameters

The estimated beta-HMM provides reduced-order summaries of the scaled spectral dynamics. Between any two states, we can compare the spectral power in a given frequency band, as well as duration statistics derived from the transition matrix.

#### Summarizing scaled spectral power in a given frequency band

The set of beta distributions provides rich information about the scaled spectral observations. One helpful statistic to summarize each beta distribution is *Pr*(*Y*_*h*_ *>* 0.5|*Z* = *k*) where (*Y*_*h*_|*Z* = *k*) ∼ *Beta*(*a*_*hk*_, *b*_*hk*_) (see glossary in Table 1). For example, when this probability is high, we can say that the scaled power for the *k*-th state and *h*-th frequency band is primarily distributed between [0.5, 1]. According to Eq. (2), this also indicates high spectral power in the *h*-th frequency band for the time-points when the state *k* is active.

**Table 1:**
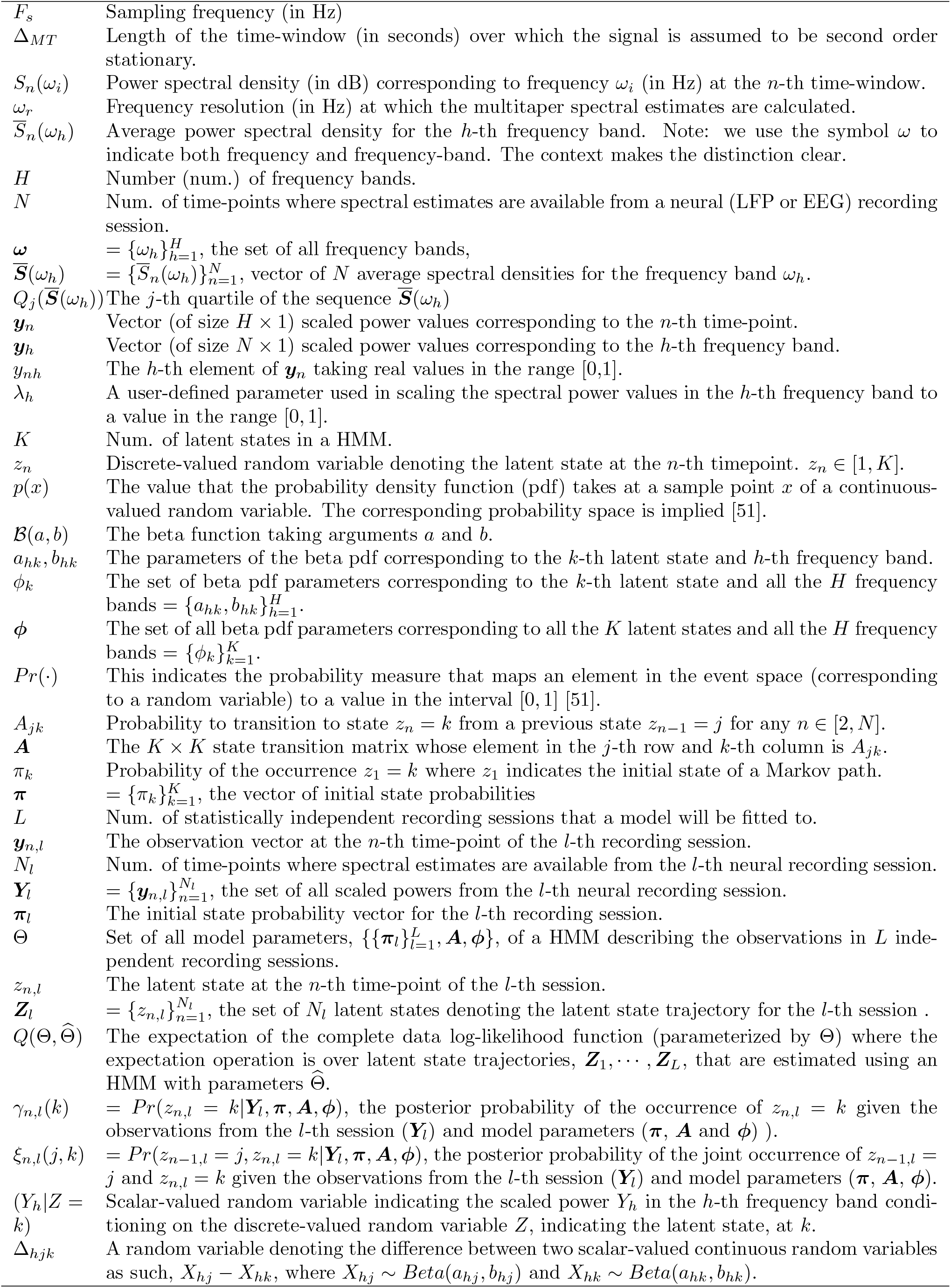
Glossary of mathematical symbols.

#### Comparing scaled spectral power in a given frequency band between any two beta-HMM states

One way to quantify how different any pair of beta distributions are from one another is to calculate the Kolmogorov-Smirnoff (KS)-distance from a large number of samples generated from the two distributions (we use 100,000 samples here). The KS-distance measures the maximum difference between two empirical CDFs, so values close to zero indicate that the underlying distributions are similar, and larger values indicate that the underlying distributions are dissimilar. Alternatively, we can leverage the analytic tractability of the beta distributions to analyze a difference random variable, Δ_*hjk*_ = *X*_*hj*_ − *X*_*hk*_, where *X*_*hj*_ ∼ *Beta*(*a*_*hj*_, *b*_*hj*_) and *X*_*hk*_ ∼ *Beta*(*a*_*hk*_, *b*_*hk*_). Here, the parameter sets {*a*_*hj*_, *b*_*hj*_} and {*a*_*hk*_, *b*_*hk*_} respectively correspond to a state *j* and another state *k* in the *h*-th frequency band. We use *Pr*(Δ_*hjk*_ ≤ 0) to denote the cumulative distribution function (cdf) value at 0 for Δ_*hjk*_ ∈ [−1, 1] such that

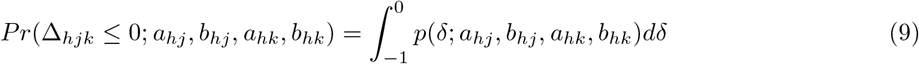

where, the pdf of the random variable Δ_*hjk*_ can be expressed at a given realization *δ* as,

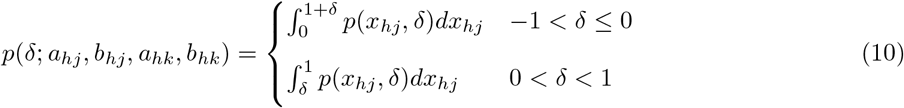

where the joint pdf *p*(*x*_*hj*_, *δ*) is defined as,

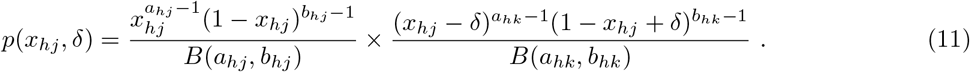

For example, if *Pr*(Δ_*hjk*_ ≤ 0) = *α*, then there is probability of *α* that *X*_*hj*_ ≤ *X*_*hk*_, or equivalently a probability of 1 − *α* that *X*_*hj*_ *> X*_*hk*_^2^. In the ensuing discussion we adopt the following notational convention for clarity. For a within-subject pairwise comparison of HMM states estimated for a single NHP (see section 4.3), we add a superscript indicating the NHP (e.g. 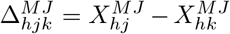, for NHP MJ). When we compare states between two NHPs, we add superscripts indicating both NHPs (e.g. 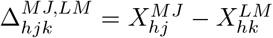, when comparing an HMM state from NHP MJ to one from NHP LM). When we perform pairwise comparison of HMM states estimated from the human OR dataset (see section 4.4), we add the superscript H, 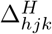.

#### Comparing duration statistics between (i) any two beta-HMM states, and (ii) any two neurophysiological states each characterized by one or more beta-HMM states

We present several statistics to describe the dynamics of the model. If *d*_*k*_ indicates a positive integer-valued random variable of the duration spent in state *k* after transitioning into it and before transitioning out to a different state, then the *mean duration* in that HMM state is given by

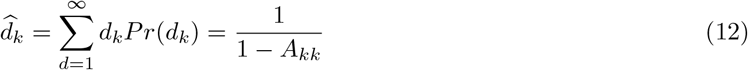

where 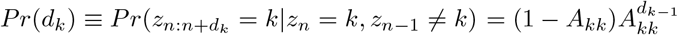, which holds for any *n >* 1 [33]. Here, the subscript (·)_,*l*_ denoting the data sequence is omitted for brevity.

In cases where a subset of beta-HMM states, ***k***^*^ ⊂ {1, …, *K*} corresponds to a neurophysiological phenomenon of interest (e.g. multiple HMM states corresponding to sub-states of gamma burst activity, or, multiple HMM states corresponding to the period between two consecutive gamma bursts), we use the following Monte Carlo approach to estimate the consecutive time spent in this subset of beta-HMM states. Using the estimated transition matrix ***Â*** and randomly sampled state *z*_1_, we generate a Markov sequence, *z*_1:*N*_ (*N* = 2000 in our study). For all *z*_*n*_’s taking values in ***k***^*^, the corresponding state label is reassigned as *z*_*n*_ = 1. The states that do not take values in ***k***^*^ are reassigned values as *z*_*n*_ = 0. We then calculate the mean duration spent in the neurophysiological state, 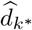, as the mean number of consecutive time points where *z*_*n*_ = 1 holds. Similarly, for the same neurophysiological state ***k***^*^ we also calculate a *mean interval* statistic as the average number of consecutive timepoints where *z*_*n*_ = 0; this is an estimate of the time interval between the two consecutive occurrences of ***k***^*^. By repeating this procedure *N*_*MC*_ times (= 4000 in our study) we calculate median and 95% confidence intervals of the state-specific mean duration and mean interval statistics, together referred to as the duration statistics in the ensuing discussion.

## 3 Experimental Recordings

### 3.1 NHP LFP recordings under ketamine anesthesia

All NHP procedures reported here followed the guidelines of the Massachusetts Institute of Technology’s Committee on Animal Care and of the National Institutes of Health. LFPs were recorded in multiple separate recording sessions from two rhesus macaques (*Macaca mulatta*) aged 14 years (NHP MJ, male, 13.0 kg) and 8 years (NHP LM, female, 6.6 kg). LFP recordings from 4 sessions in NHP MJ and 5 sessions in NHP LM were analyzed in this study. During each recording session, ketamine was administered as a single 20 mg/kg bolus intramuscular dose. This is a putative high dose for sedation and low dose for surgical anesthesia in NHPs [61, 62, 63]. Fifteen minutes prior to ketamine administration, glycopyrrolate (0.01 mg/kg) was delivered to reduce salivation and airway secretions. In both NHPs, LFPs were recordered from a 8 × 8 iridium-oxide contact microelectrode array (“Utah array”, MultiPort: 1.0 mm shank length, 400 *µ*m spacing, Blackrock Microsystems, Salt Lake City, UT) implanted in the prefrontal cortex (area 8A). LFPs, recorded at 30 kHz, were low-pass filtered to 250 Hz and then downsampled to 1 kHz. LFPs were continuously recorded from 1−5 minutes prior to ketamine injection up to 18−20 minutes following ketamine injection. In the ensuing figures representing neural timeseries data, the time 0 denotes the time-point when the ketamine bolus was administered.

### 3.2 Human EEG recordings under ketamine anesthesia

The Partners Institutional Review Board approved this human retrospective observational study. The data analyzed in this manuscript were acquired during an EEG study of ketamine-induced general anesthesia. We reviewed and selected 9 patients (Age: 51.8 *±* 11.7 years, Weight: 84.5 *±* 15.5 kg, mean *±* standard deviation) from our database who were administered an intravenous bolus dose of ketamine as the sole hypnotic drug for the induction of general anesthesia. The ASA scores in all these 9 patients were less than or equal to 3, and none of the patients had a history of any neurological or psychiatric disorders. Prior to the ketamine bolus, patients were administered midazolam (n=8; 1.81 *±* 0.53 mg) and/or fentanyl (n=7; 164.29 *±* 80.18 mcg) for anxiolysis and to block the sympathetic response to laryngoscopy, respectively. To induce general anesthesia (GA) a bolus dose of ketamine (mean *±* standard deviation; 182.22 *±* 29.06 mg, 10 mg/ml) was administered, and intubation was carried out using succinylcholine, cisatracurium, or rocuronium for muscle relaxation. EEG data were acquired using Sedline monitor (Masimo Inc, USA). The standard Sedline Sedtrace electrode arrays were placed on the forehead that approximated the positions of Fp1, Fp2, F7, and F8, the ground electrode at Fpz, and the reference electrode 1 cm above Fpz. For our analysis, we use the data recorded from Fp2. Data were recorded with a pre-amplifier bandwidth of 0.5 to 92 Hz, a sampling rate of 250 Hz, with 16-bit, 29 nV resolution. Electrode impedance was less than 5 kΩ in each channel. We analyzed continuous EEG recordings starting from 2 minutes pre-ketamine bolus and up to 5.5 min - 14 min post-ketamine bolus, and prior to the administration of any additional hypnotic drugs used to maintain GA.

## 4 Results

### 4.1 Testing the beta-HMM analysis framework against simulated ground truth

To test the ability of our beta-HMM analysis framework to accurately retrieve a known Markov sequence and associated observation distributions, we simulated spectrograms generated by pre-specified Markov processes (See Table 2 in the appendix for simulation algorithm). We simulated spectral dynamics comparable to those caused by ketamine by using a spectrogram *S*_*n*_(·) calculated from one session of NHP experimental data (as described in Sec 2.1) as input to the algorithm (Table 2 in the appendix). Additional inputs to this algorithm are user-prescribed HMM parameters, the number of states *K*, ***π*** and ***A***, as well as the duration *M* of the simulated data. This algorithm outputs a realization of the latent state path, 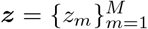, corresponding simulated spectrogram, 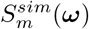, and ***Y***. While 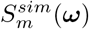 is used for visualization, the realization of ***Y*** serves as the observations from which we estimate a *K*-state beta-HMM (Sec. 2.4). Salient features of the algorithm in Table 2 of the appendix are as follows. First, *K* distinct spectral clusters are created from the given NHP LFP spectrogram using an unsupervised clustering algorithm (different from beta-HMM) that does not respect the dependence across consecutive time-points. Then a Markov sequence is simulated using the known parameters, ***π*** and ***A***. Finally, to generate the spectrogram with *K* underlying latent states with Markov state transitions, a spectrum is selected for a given instant by uniformly sampling from one of the *K* spectral clusters that corresponds to the realization of the latent state at the same instant.

**Table 2:** Algorithm to simulate data from an NHP LFP session with an underlying Markovian latent state path for numerically testing the beta-HMM framework

|  |  |
| --- | --- |
| **Input** | Spectrogram $\{S_n(\omega_i)\}_{n=1}^N$ (for every frequency $\omega_i \in \{\omega_r, 2\omega_r, \dots, 50\}$) from one LFP recording session from NHP MJ of duration 20 min, user-specified number of states $K$ and HMM parameters $\boldsymbol{\pi}$ and $\mathbf{A}$, and length of session to simulate, $M = 12000$. |
| 1 | Calculate $\{\bar{S}_n(\boldsymbol{\omega})\}_{n=1}^N$ (Eq. 1). |
| 2 | Identify $K$ distinct clusters of spectra by executing a k-means clustering algorithm ([31]) on $\{\bar{S}_n(\boldsymbol{\omega})\}_{n=1}^N$. Label each cluster from 1 to $K$ in ascending order of within-cluster mean power in the 40-50 Hz frequency band. Note: Each time-point $n$ is now associated with a cluster label 1 to $K$. |
| 3 | Store the original time-point indices corresponding to each element in $k$-th cluster in an integer-valued set $\mathbf{n}_k$, where $k \in [1, K]$. Note: $\bigcup_{k=1}^K \mathbf{n}_k = [1, N]$ and $\bigcap_{k=1}^K \mathbf{n}_k = \emptyset$, where ∅ denotes the null set. |
| 4 | Simulate a state path by generating a Markov sequence $\{z_m\}_{m=1}^M$ using the set of parameters $\{\boldsymbol{\pi}, \mathbf{A}\}$. |
| 5 | Simulate an LFP spectrogram, $\{S_m^{sim}(\cdot)\}_{m=1}^M$, as follows: when $z_m = k$, uniformly sample index $n_k$ from $\mathbf{n}_k$, and set $S_m^{sim}(\omega_i) = S_{n_k}(\omega_i)$ for every frequency $\omega_i \in \{\omega_r, 2\omega_r, \dots, 50\}$. Also, set the frequency band-specific average of the simulated spectrum corresponding to $z_m = k$, $\bar{S}_m^{sim}(\boldsymbol{\omega}) = \bar{S}_{n_k}(\boldsymbol{\omega})$. |
| 6 | Calculate $\mathbf{Y} = \{\mathbf{y}_m\}_{m=1}^M$ using $\{\bar{S}_m^{sim}(\boldsymbol{\omega})\}_{m=1}^M$ (Eq. 2). |
| 7 | Determine cluster-specific and frequency-band specific maximum-likelihood estimate of the beta distribution parameters, $\{a_{hk}, b_{hk}\}$, for all $h = \{1, 2, \dots, H\}$ and $k = \{1, 2, \dots, K\}$ from $\mathbf{Y}$. Note: $\boldsymbol{\phi} = \{\{a_{hk}, b_{hk}\}_{h=1}^H\}_{k=1}^K$ estimated based on the simulated state path is considered as *ground truth* for the purpose of numerically testing the beta-HMM estimation framework. |
| **Output** | $\{S_m^{sim}(\omega_i)\}_{m=1}^M$ (for every frequency $\omega_i \in \{\omega_r, 2\omega_r, \dots, 50\}$) and corresponding $\mathbf{Y}$, $\{z_m\}_{m=1}^M$, and $\boldsymbol{\phi}$. |

For a given *K*, we generate 100 realizations of ***Y*** based on *K* spectral clusters. For each realization of ***Y***, we estimate a *K*-state beta HMM to determine a set of estimated model parameters 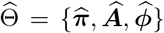, and the estimated state path ***z***^*^ (calculated using the Viterbi algorithm). We assess goodness of fit for each realization of ***Y*** by comparing the estimated state path, ***z***^*^, and model parameters, 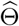, to the ground truth state path, ***z***, and model parameters, Θ, respectively.

To assess the accuracy of the estimated path ***z***^*^ compared to the known path ***z***, we calculated the fraction of time points for which the estimated and known path are identical:

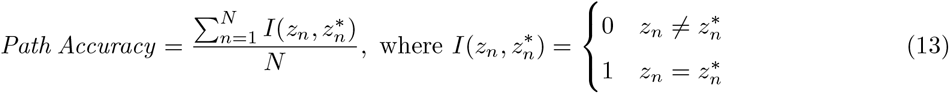

To assess the accuracy of the observation distribution estimation, we calculated the mean KS-distance between the *HK* ground truth and estimated beta distributions. KS-distances close to zero indicate that the estimated and ground truth distributions are similar. We also calculated the absolute errors in the estimated initial state and state transition probabilities, denoted respectively by *ϵ*_*A*_ and *ϵ*_*π*_,

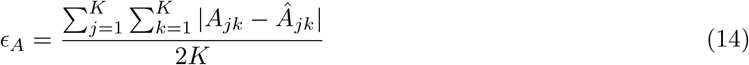

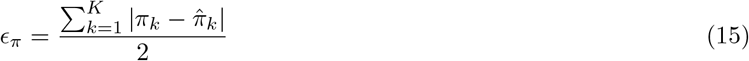

Note the the maximum possible value for 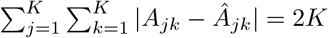 and for 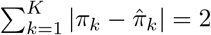. Thus, the denominators scale the absolute errors to the range of [0, 1]. We report the median and 90% confidence intervals of each of these metrics calculated across 100 simulated spectrograms for each model order in Fig. 2.

**Figure 2:**
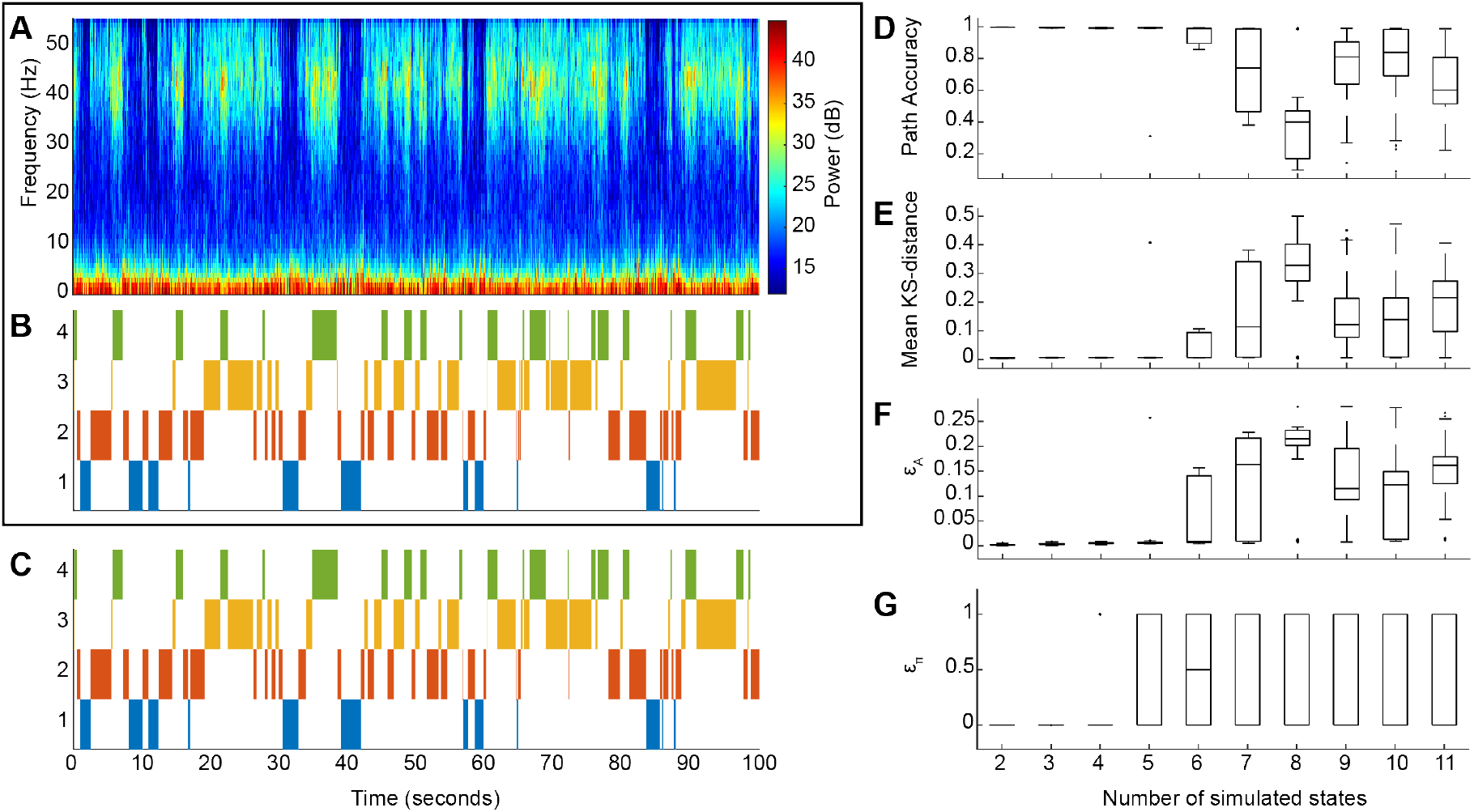
The beta-HMM tracks the dynamic structure of simulated spectrograms characterized by discrete state transitions. (extracted from one sequence of ketamine-induced LFP in NHP MJ). (A) One realization of simulated spectrogram. (B) Markov path corresponding to the simulated spectrogram in panel A. (C) Optimal state trajectory estimated from simulated data in panel A via the beta-HMM analysis. (D-G) Goodness of fit analysis. Box-plots summarize the state path accuracy (Eq. (13)) in panel D, the KS-distance between the estimated and the actual state-specific and frequency band-specific beta distributions (averaged across all states and all frequency bands) in panel E, the relative error in transition matrix (Eq. (14)) in panel F, and relative error in the initial state probabilities (Eq. (15)) in panel G. For each specified number of states, 100 realizations of spectrograms were simulated and the beta-HMM analysis was performed on each.

Simulation analysis revealed that the model parameters of simulated spectrograms, generated from ketamine-induced neural data and characterized by known Markov transition dynamics, can be accurately estimated with the beta-HMM framework for low model orders. The performance of the beta-HMM analysis framework on a typical simulated spectrogram (Fig. 2(A)) demonstrates that the estimated state trajectory (Fig. 2(C)) matches well with the actual state trajectory (Fig. 2(B)). Across 400 simulated datasets for models with 2 through 5 states (100 simulations each), the estimated state trajectory was estimated with high accuracy (*Path Accuracy >* 0.98 for all but one of 400 independent simulations), the beta distributions were estimated with high accuracy (mean KS-distance *<* 7.76 × 10^−3^ for all but one of 400 independent simulations) and the transition matrix was estimated with low relative error (*ϵ*_*A*_ *<* 0.01 for all but one of 400 independent simulations). For models with 2-3 states, the initial probability, which corresponds to a single data point, was also accurately estimated (*ϵ*_*π*_ *<* 2.45 × 10^−14^). Beyond 5 states, there was a significant drop-off in estimation accuracy. A potential reason for this occurrence is redundancy across some of the *K >* 5 spectral clusters derived from the original NHP spectrogram, which may not contain more than 5 clusters of distinct neural activity. Overall, this numerical experiment generated detailed insights on the level of inaccuracy in our beta-HMMs when estimated from spectral data derived from real neural activity induced by ketamine.

### 4.2 Demonstration of the beta-HMM analysis on a single observation sequence of NHP LFP data (*L* = 1 case)

Using the numerically tested beta-HMM analysis framework, we analyzed a single session high-dose ketamine NHP LFP recording (Sec 3). From qualitative analysis of the time-series data and as well as spectral estimates (Fig. 9 in Supporting Figures section), we identified 4 key signatures induced by ketamine: a *slow oscillation state* with a prominent low-frequency (0-10 Hz) activity and negligible activity in the gamma band (30-50 Hz); a transition state with simultaneous activity in 0-10 Hz and 30-50 Hz bands; and two others with significant but differing levels of gamma activity and relatively lower low-frequency activity, which we refer to as *gamma burst states*. We also sought to capture the transition from pre- to post-ketamine bolus neurophysiology, so we chose a model order of *K* = 5. We estimated a 5-state beta-HMM from a single sequence of LFP spectrogram.

**Figure 3:**
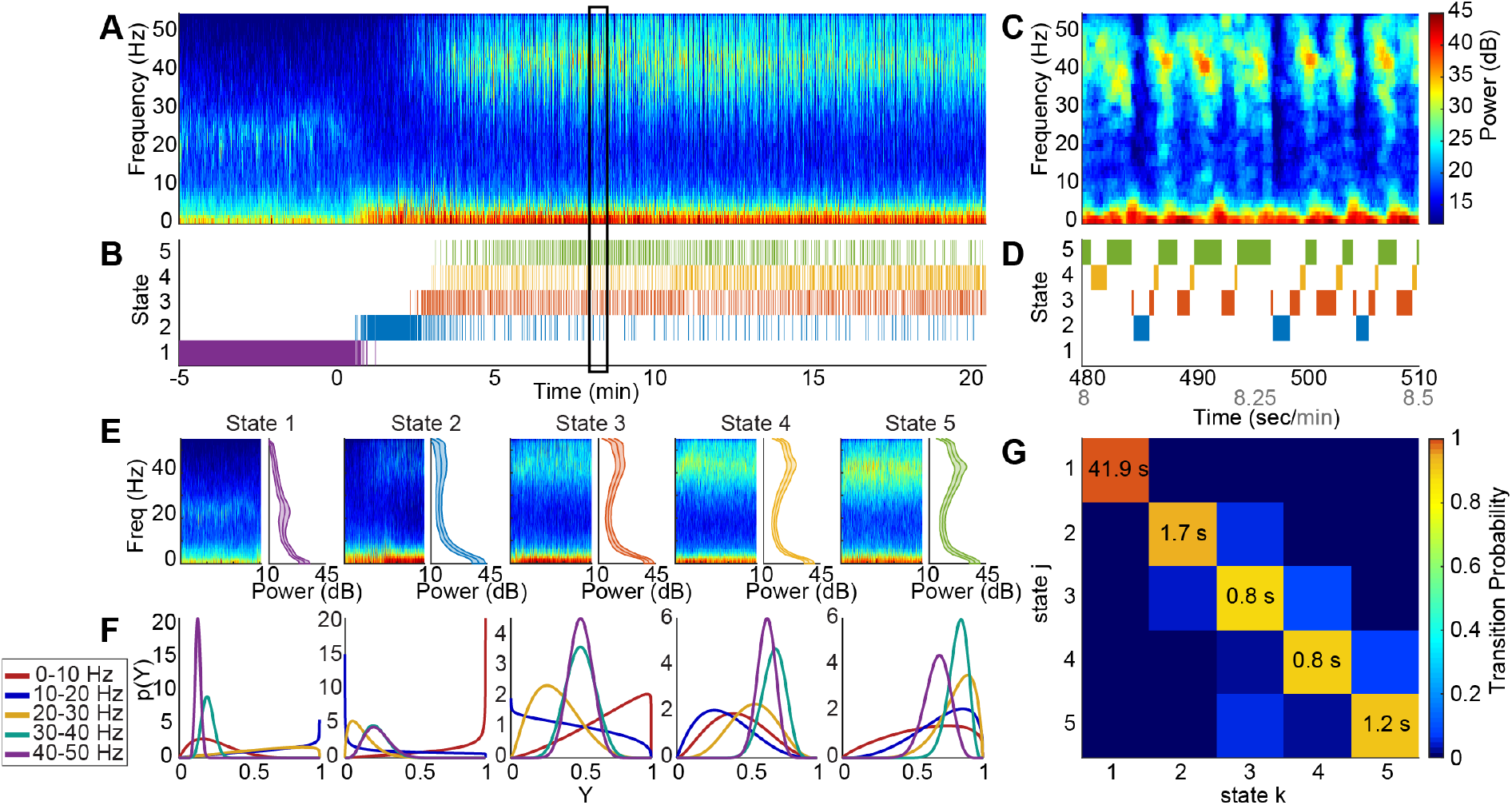
A 5-state beta-HMM estimates ketamine-induced neurophysiological dynamics from a single NHP LFP sequence. (A) Multitaper spectrogram of LFP where time 0 marks the time-point of administration of 20 mg/kg ketamine intramuscular bolus. (B) Estimated latent state trajectory. (C) Enlarged view of panel A for the epoch identified by the rectangular box. (D) Enlarged view of panel B for the epoch identified by the rectangular box. (E) Spectral clusters identified by the 5-state beta-HMM. Each spectral cluster has a characteristic signature as indicated by the mean spectrum and corresponding standard deviation. (F) Frequency-band specific beta distributions (over scaled power (Y) in respective bands) estimated for each of the 5 states. (G) The estimated transition matrix. The expected value of the duration spent in each state is indicated on the diagonal.

**Figure 4:**
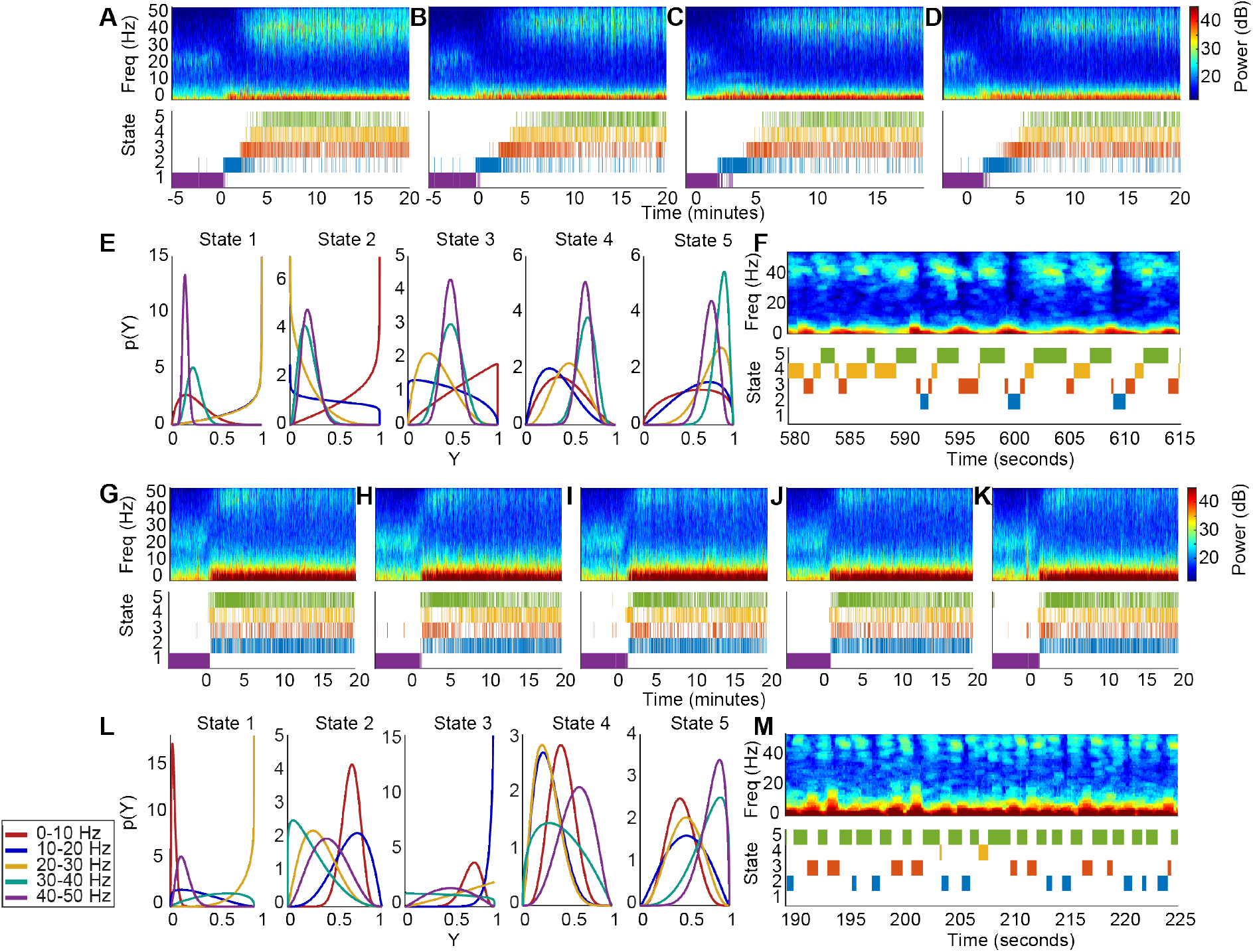
The 5-state beta-HMM can be fit across multiple sessions to create an NHP-specific model. (A-D) Multitaper spectrograms of LFP and corresponding estimated latent state trajectories from 4 sessions in NHP MJ. (E) Frequency band-specific beta pdfs for each of the 5 states estimated from the 4 sessions in NHP MJ (Y - scaled power) (F) Enlarged view of a typical 35s epoch in panel D. (G-K) Multitaper spectrograms of LFP and corresponding estimated latent state trajectories from 5 sessions in NHP LM. (L) Frequency band-specific beta pdfs for each of the 5 states estimated from the 5 sessions in NHP LM. (Y - scaled power) (M) Enlarged view of a typical 35s epoch in panel K.

**Figure 5:**
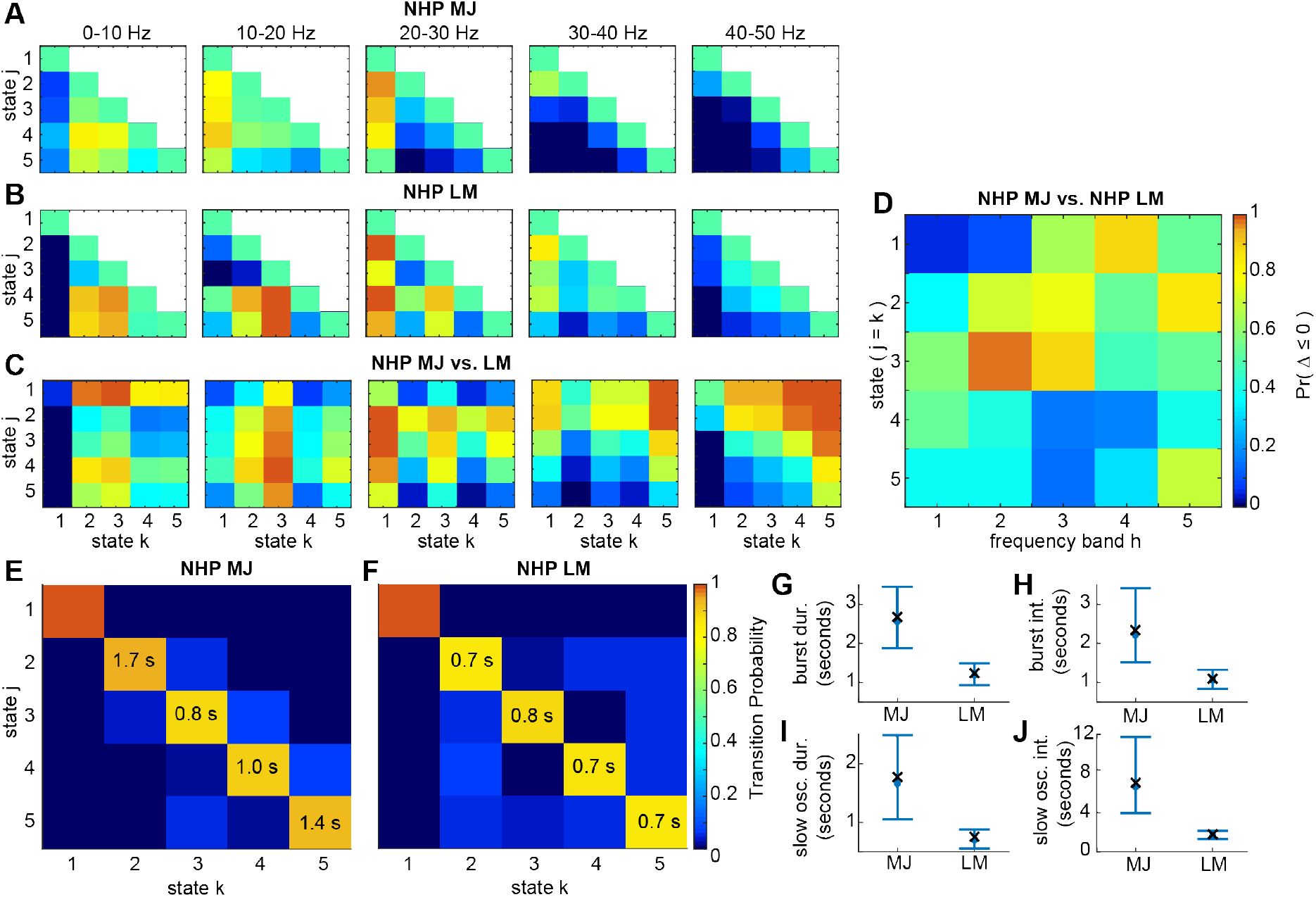
The estimated beta pdfs and transition matrices characterize the underlying dynamics of NHP LFP data following a 20 mg/kg bolus of ketamine. (A-C) Heatmaps indicating probability of the event that scaled power in state *j* (of NHP MJ in panels A and C, and of NHP LM in panel B) is less than or equal to the scaled power in state *k* (of NHP MJ in panel A, and of NHP LM in panels B and C) for 1 ≤ *k* ≤ *j* ≤ 5 and for each of the 5 frequency bands. The color indicates *Pr*(Δ ≤ 0) which represents *Pr*(*X*_*hj*_ − *X*_*hk*_ ≤ 0), where *X*_*hj*_ is a random variable characterized by state *j*’s beta pdf in frequency band *h* and *X*_*hk*_ is a random variable characterized by state *k*’s beta pdf in the same frequency band *h*. (D) Heatmap representing diagonal elements from each of the matrices in panel C rearranged columnwise to create a 5 × 5 matrix wherein a row indicates a state pair (1 ≤ *j* = *k* ≤ 5) compared between NHP MJ and NHP LM, and a column indicates a frequency band. (E, F) The transition matrices for NHP MJ and NHP LM with expected state duration (in seconds) indicated on the diagonal for all states except the first state. (G-J) Duration statistics corresponding to gamma burst and slow oscillation states in NHP MJ and NHP LM. For each subject, 4000 Markov sequences of length *N* = 2000 were simulated using the subject-specific estimated transition matrix. The mean duration and interval corresponding to the gamma burst and slow oscillation states were calculated for each realization of these sequences. Median and 95% confidence bounds across all the subject-specific simulated sequences are indicated in blue. The mean duration and interval calculated from the estimated state trajectory (output of the Viterbi algorithm which uses the maximum likelihood beta-HMM parameters and the observations as input) are indicated by a cross (**×**) symbol.

**Figure 6:**
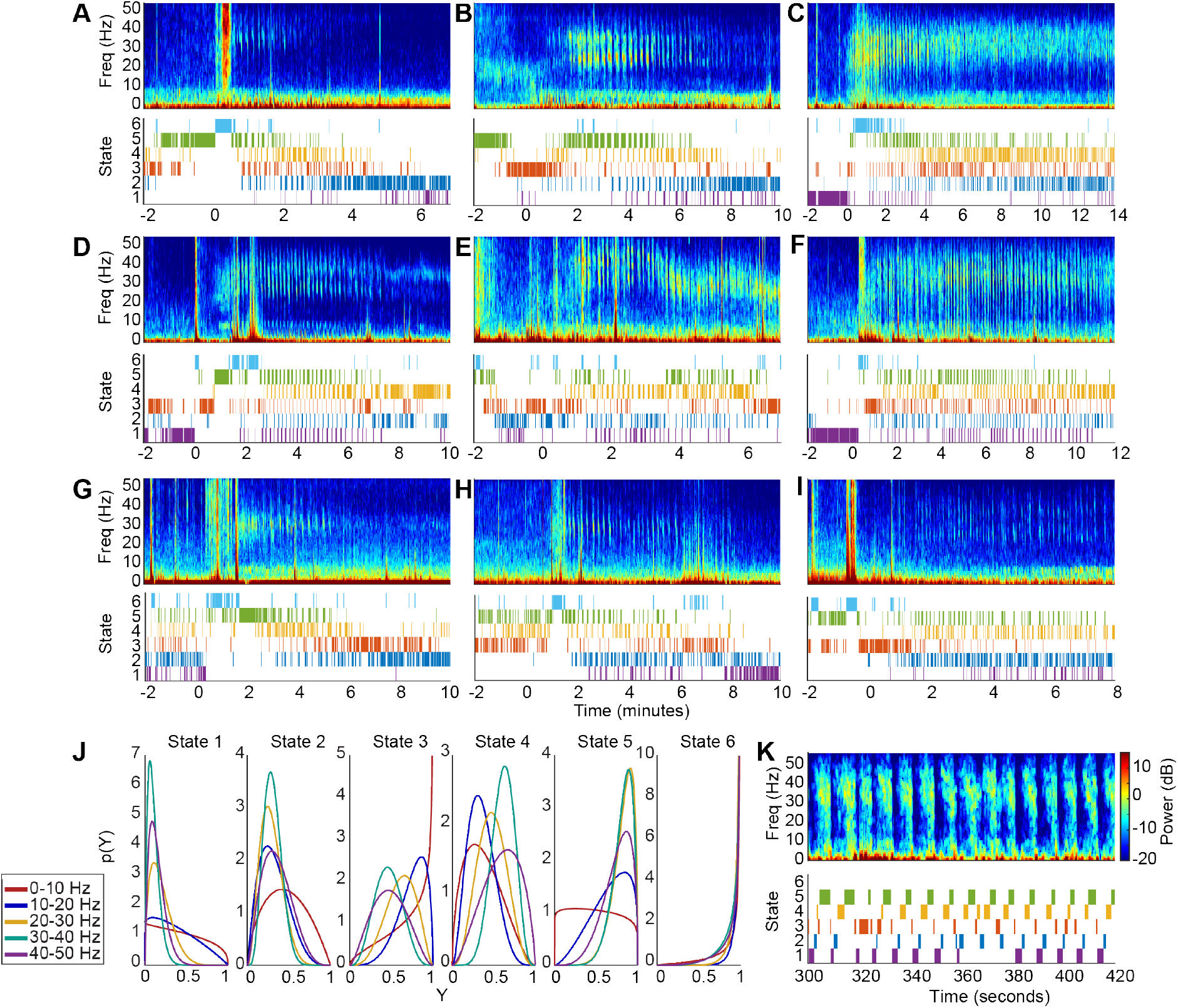
The 6-state beta-HMM can be fit across independent EEG recordings from multiple human subjects to create a typical human-specific model. (A-I) Multitaper spectrograms of EEG and corresponding estimated latent state trajectories from 9 human subjects. (J) Frequency band-specific beta pdfs for each of the 6 states estimated from the 9 EEG sessions. (Y - scaled power) (K) Enlarged view of a typical 120s epoch in panel F.

**Figure 7:**
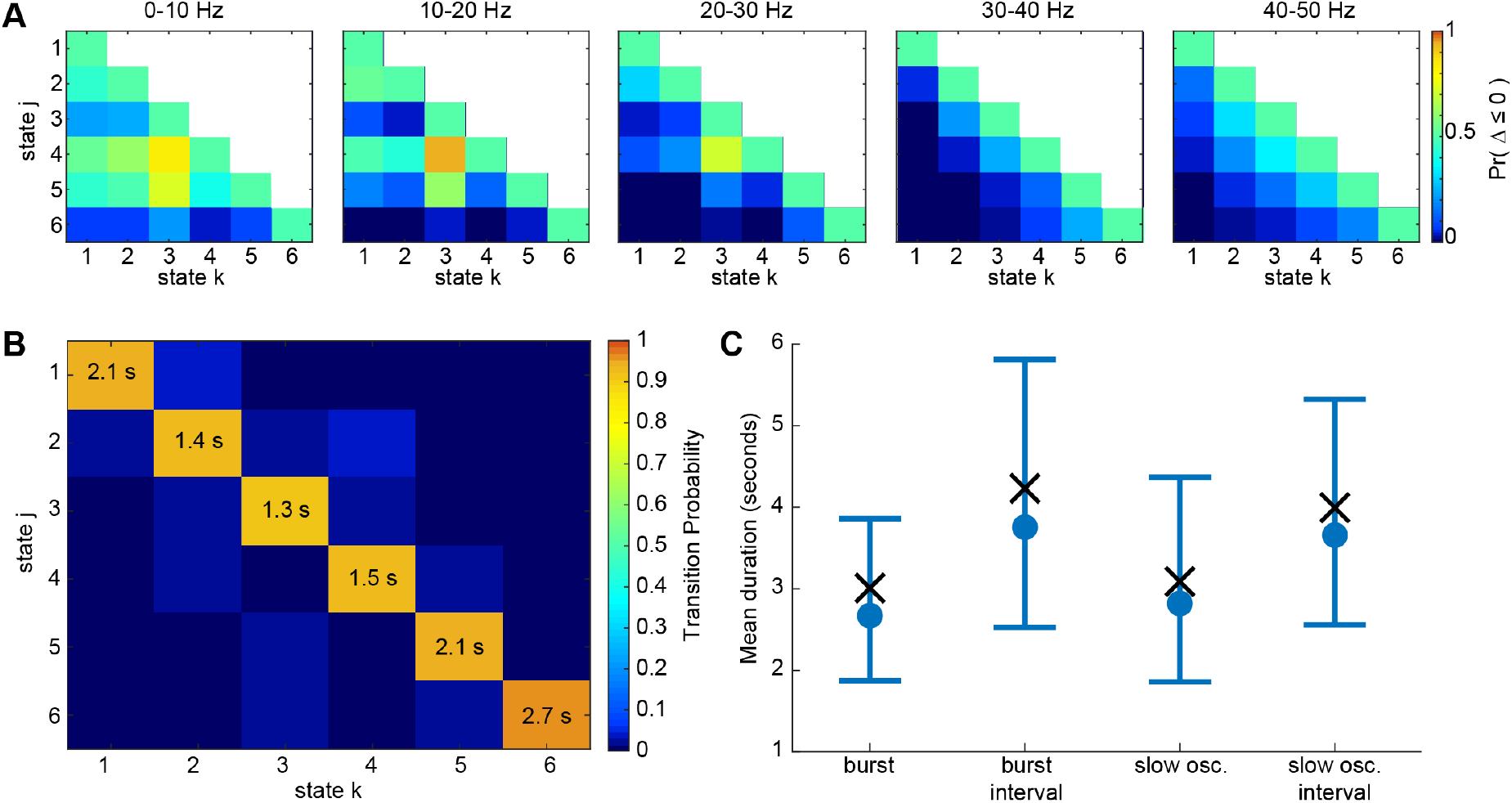
The estimated beta pdfs and transition matrix characterize the underlying dynamics of human scalp EEG under ketamine-induced general anesthesia. (A) Heatmaps indicating probability of the event that scaled power in state *j* is less than or equal to the scaled power in state *k* for 1 ≤ *k* ≤ *j* ≤ 6 and for each of the 5 frequency bands. The color indicates *Pr*(Δ ≤ 0) which represents *Pr*(*X*_*hj*_ − *X*_*hk*_ ≤ 0), where *X*_*hj*_ is a random variable characterized by state *j*’s beta pdf in frequency band *h* and *X*_*hk*_ is a random variable characterized by state *k*’s beta pdf in the same frequency band *h*. (B) The transition matrices for NHP MJ and NHP LM with expected state duration (in seconds) indicated on the diagonal for all states. (C) Duration statistics corresponding to gamma burst and slow oscillation states. 4000 Markov sequences of length *N* = 2000 were simulated using the estimated transition matrix. The mean duration and interval corresponding to the gamma burst and slow oscillation states were calculated for each realization of these sequences. Median and 95% confidence bounds across all the simulated sequences are indicated in blue. The mean duration and interval calculated from the estimated state trajectory (output of the Viterbi algorithm which uses the maximum likelihood beta-HMM parameters and the observations as input) are indicated by a cross (**×**) symbol.

**Figure 8:**
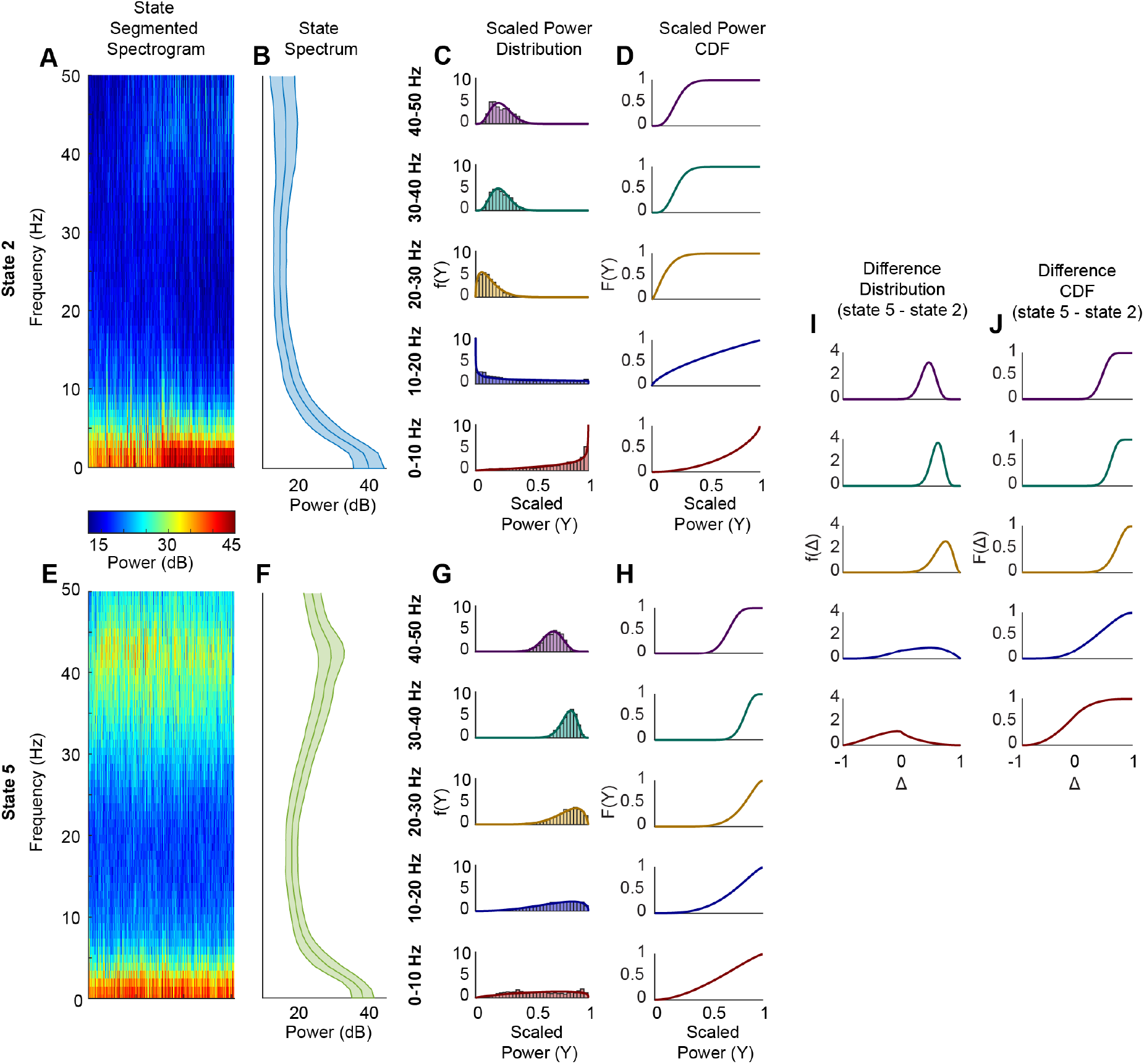
Illustrative example of how to interpret the state-specific spectral profiles in terms of the state-specific frequency-band specific beta pdf’s. (A, E) Group of spectra from time-points identified as states 2 and 5 (by the Viterbi algorithm), respectively, of a 5 state beta-HMM fitted to a single session of NHP data. (B,F) The average and standard deviation across all the spectral profiles for states 2 and 5, respectively. (C, G) Empirical pdfs of the scaled power in the 5 frequency bands, based on the data from the time-points identified as states 2 and 5 (by the Viterbi algorithm), respectively. The state-specific frequency-band specific beta pdfs of the maximum likelihood beta-HMM (estimated by the EM algorithm) is overlaid. (D, H) The cdf corresponding to the estimated beta pdfs plotted in C and G, respectively. (I) The frequency-band specific pdf of the difference random variable Δ = *X*_*h*,5_ − *X*_*h*,2_, where *X*_*h*,5_ ∼ *Beta*(*a*_*h*,5_, *b*_*h*,5_) and *X*_5,*h*_ ∼*Beta*(*a*_*h*,2_, *b*_*h*,2_) based on the respective estimated pdfs, (J) The cdf corresponding to the frequency band-specific pdf’s in (I).

**Figure 9:**
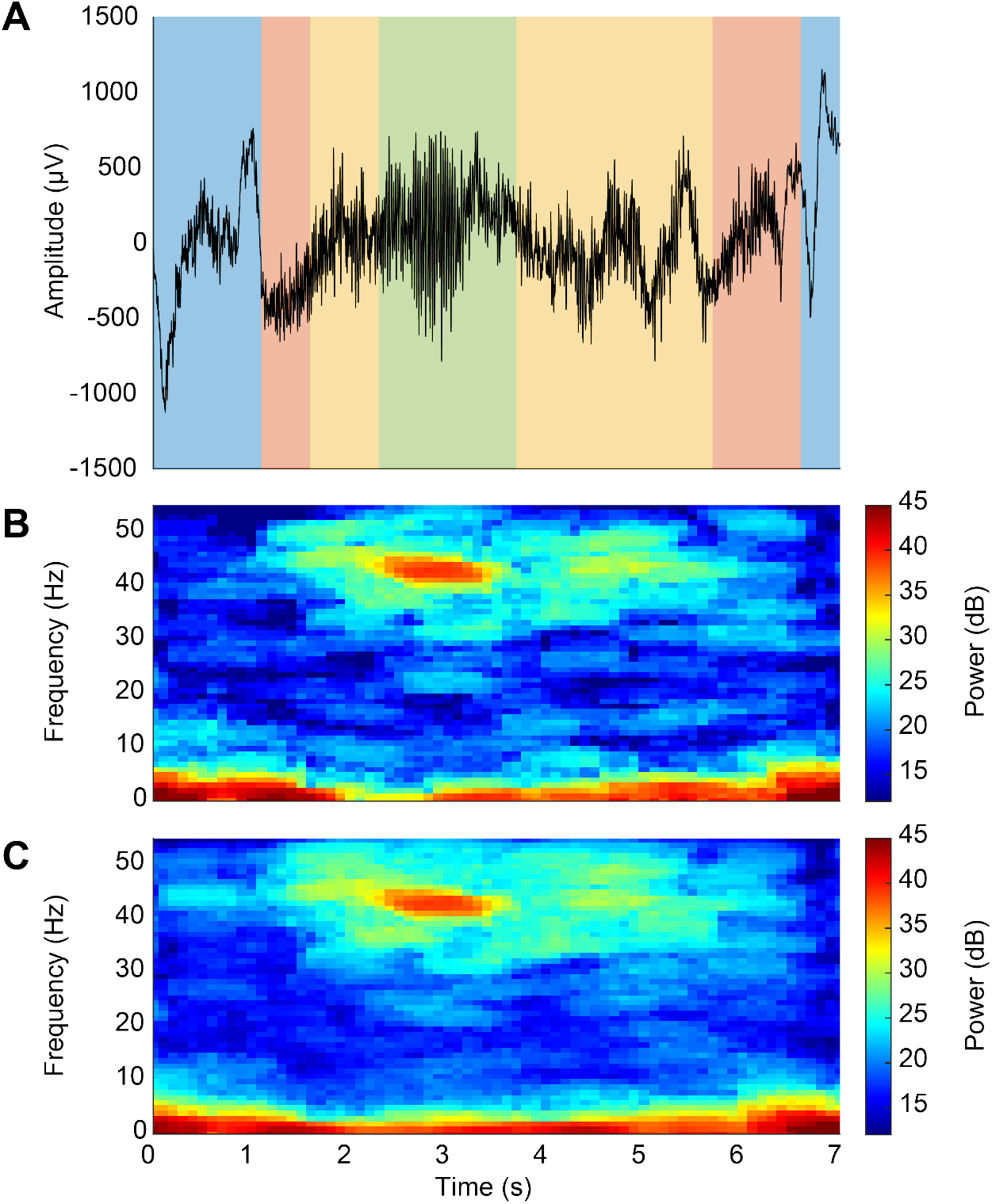
A typical 7 second epoch (following a high-dose ketamine bolus) of LFP from a single electrode of a multi-electrode array located in the PFC (area 8A) of NHP MJ is presented in panel A. Time 0 corresponds to 743.5 seconds after the bolus was administered. We fit our beta-HMM to the reduced-order representation (derived using Eqs (1) and (2)) of the corresponding multitaper spectrogram, presented in panel B. The resulting optimal segmentation is represented by the colored vertical bars overlaid on the neural time-series in panel A. State 2 is shown in blue, state 3 in red, state 4 in yellow, and state 5 in green. (State 1, which corresponds to the time before the ketamine bolus, is not shown here.) Panel C represents the average spectrogram across all the electrodes in the same multi-electrode array.

The 5-state beta-HMM provided a quantitative characterization of the neurophysiological dynamics induced by ketamine (see Fig. 3(A, B), as well as Fig. 10 in the Supporting Figures section). The beta-HMM analysis objectively identified gamma-slow dynamics in NHPs similar to those described by [11]. We found that the HMM identified alternating states which corresponded to periods of prominent gamma oscillations, prominent slow oscillations in the absence of gamma, and an intermediate state in between (Fig. 3(C, D)). From visual inspection of the the state-segmented spectrograms and further analysis of beta distributions (Fig. 3(E, F)), we identified the relevant neurophysiological states with distinct spectral signatures. For example, in the state-segmented spectrogram for state 5 (Figure 3E), we can see that high spectral power in the 30-40 Hz frequency band (h = 4) is associated with a high probability that (*Y*_4_|*Z* = 5) *>* 0.5 (Figure 3F)^3^. Based on the beta distributions, we found that HMM states 4 and 5 both have high gamma power (30-40 Hz (h = 4): *Pr*(*Y*_4_ *>* 0.5|*Z* = 4) = 0.98, *Pr*(*Y*_4_ *>* 0.5|*Z* = 5) = 1.00; 40-50 Hz (h = 5): *Pr*(*Y*_5_ *>* 0.5|*Z* = 4) = 0.98, *Pr*(*Y*_5_ *>* 0.5|*Z* = 5) = 0.97). HMM state 2 is dominated by prominent slow power (0-10 Hz (h = 1): *Pr*(*Y*_1_ *>* 0.5|*Z* = 2) = 0.82) and low gamma power (*Pr*(*Y*_4_ *>* 0.5|*Z* = 2) = 0.00, *Pr*(*Y*_5_ *>* 0.5|*Z* = 2) = 0.00). Therefore, we consider HMM states 4 and 5 together to represent the neurophysiological gamma burst state, HMM state 2 to represent the slow oscillation state, and HMM state 3 to represent the transition between neurophysiological slow oscillation and gamma burst states.

**Figure 10:**
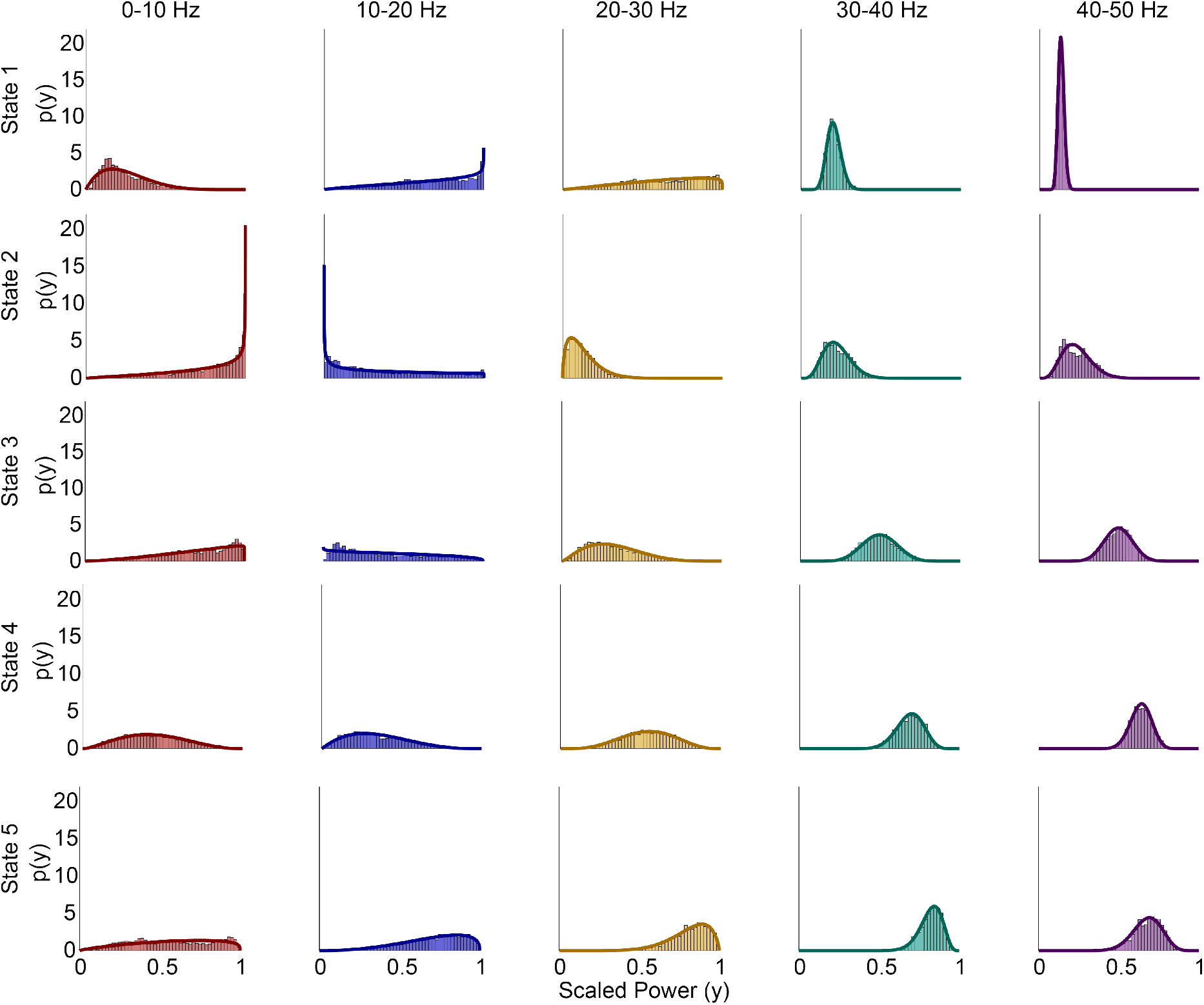
Illustration of the state-specific and frequency-band specific beta pdfs corresponding to a 5-state beta-HMM that is estimated from *L* = 1 session of LFP recording from NHP MJ. For each state (viewed row-wise) and a frequency band (viewed column-wise), the corresponding subplot presents (1) the empirical pdf plotted as a histogram based on the observations that correspond to the optimal segmentation of the data sequence, and (2) the continuous beta pdf with parameters estimated by the EM algorithm. Each color distinguishes a frequency band.

The transition matrix (Fig. 3G) quantifies the alternating dynamics noted in Fig. 3(C, D). From the transition matrix, we can also determine the expected duration of each state, as denoted on the diagonal terms indicating self-transition probabilities (Eq. 12). From the transition probabilities and optimal state trajectory, we infer there is a high probability of the state sequence 2 → 3 → 4 → 5 → 3 → 2. This confirms the alternating dynamics between time-localized high power activity in the gamma and low frequency bands induced by ketamine. The beta-HMM provides further evidence that the neural activity induced by ketamine is distinct from pre-ketamine neural activity. States 2 through 5 do not occur prior to the ketamine bolus (Fig. 3(A, B)). Furthermore, HMM state 1 has sustained prevalence prior to the bolus and does not occur after a brief initial period (approximately 1 minute) post-bolus. Therefore, HMM state 1 can be associated with a pre-ketamine neurophysiological state. This is further supported by the fact that the transition probability from any of the post-bolus states back to state 1 is less than 0.003.

### 4.3 Subject-specific beta-HMMs estimated from multiple recording sessions (*L >* 1 case) from 2 NHPs

A review of summary statistics from session-specific beta-HMMs estimated from multiple recording sessions of each NHP indicated the presence of subject-specific HMM states that have common features (both spectral and temporal) across sessions. This further suggested the utility of a subject-specific HMM for each NHP. Therefore, we fitted two 5-state beta-HMMs to multiple LFP recording sessions from 2 NHPs: 4 sessions for NHP MJ and 5 sessions for NHP LM. A key implementation feature in our beta HMM analysis from multiple sessions (for a given group) is that we do not concatenate multiple sessions; rather we treat them as mutually independent in our EM algorithm implementation (as discussed in Sec. 2.4). Thus, the beta-HMM analysis respects the sequential nature of the observations within a session, while maintaining the distinction across multiple sessions. Since there were at least three days between each NHP session, we treated them as mutually independent and therefore utilized the EM algorithm based on Eq. (8) to estimate the model parameters.

We present the optimal state trajectories and state-specific and frequency band-specific observation distributions estimated from all 9 sessions of 2 NHPs in Fig. 4 (also see Figs. 11 and 12 in the Supporting Figures section). Although the amplitude of the spectral power tended to vary across sessions, our use of scaled power facilitated identification of common states with similar spectral profiles across multiple sessions. In both NHPs, we found that the beta distributions in each frequency band were unique across all states (KS-distance *>* 0.05 for all pairwise comparisons; 100 comparisons = 10 pairs of states per frequency band per NHP × 5 frequency bands × 2 NHPs) (Fig. 4(E,L)). Consistent with the single session observations in NHP MJ, in both primates we found that HMM state 5 has high scaled power in the 30-40 Hz range, and both HMM states 4 and 5 have high power in the 40-50 Hz range (NHP MJ: *Pr*(*Y*_4_ *>* 0.5|*Z* = 5) = 1.00, *Pr*(*Y*_5_ *>* 0.5|*Z* = 4) = 0.97, *Pr*(*Y*_5_ *>* 0.5|*Z* = 5) = 0.99; NHP LM: *Pr*(*Y*_4_ *>* 0.5|*Z* = 5) = 0.90, *Pr*(*Y*_5_ *>* 0.5|*Z* = 4) = 0.71, *Pr*(*Y*_5_ *>* 0.5|*Z* = 5) = 0.97)^4^. We found that state 4 in NHP LM had comparatively lower scaled power in the 30-40 Hz range (NHP MJ: *Pr*(*Y*_4_ *>* 0.5|*Z* = 4) = 0.95; NHP LM: *Pr*(*Y*_4_ *>* 0.5|*Z* = 4) = 0.36). We also found in both NHPs that state 2 is dominated by prominent slow power (NHP MJ: *Pr*(*Y*_1_ *>* 0.5|*Z* = 2) = 0.81; NHP LM: *Pr*(*Y*_1_ *>* 0.5|*Z* = 2) = 0.96) and low gamma power (NHP MJ: *Pr*(*Y*_4_ *>* 0.5|*Z* = 2) = 0.01, *Pr*(*Y*_5_ *>* 0.5|*Z* = 2) = 0.00; NHP LM: *Pr*(*Y*_4_ *>* 0.5|*Z* = 2) = 0.12, *Pr*(*Y*_5_ *>* 0.5|*Z* = 2) = 0.37). Again, we consider HMM states 4 and 5 together to represent the neurophysiological gamma burst state, and HMM state 2 to represent the slow oscillation state. As demonstrated in Fig. 3(C,D), the multisession beta-HMM model captures the alternating dynamics induced by ketamine in both primates (Fig. 4(F,M)).

**Figure 11:**
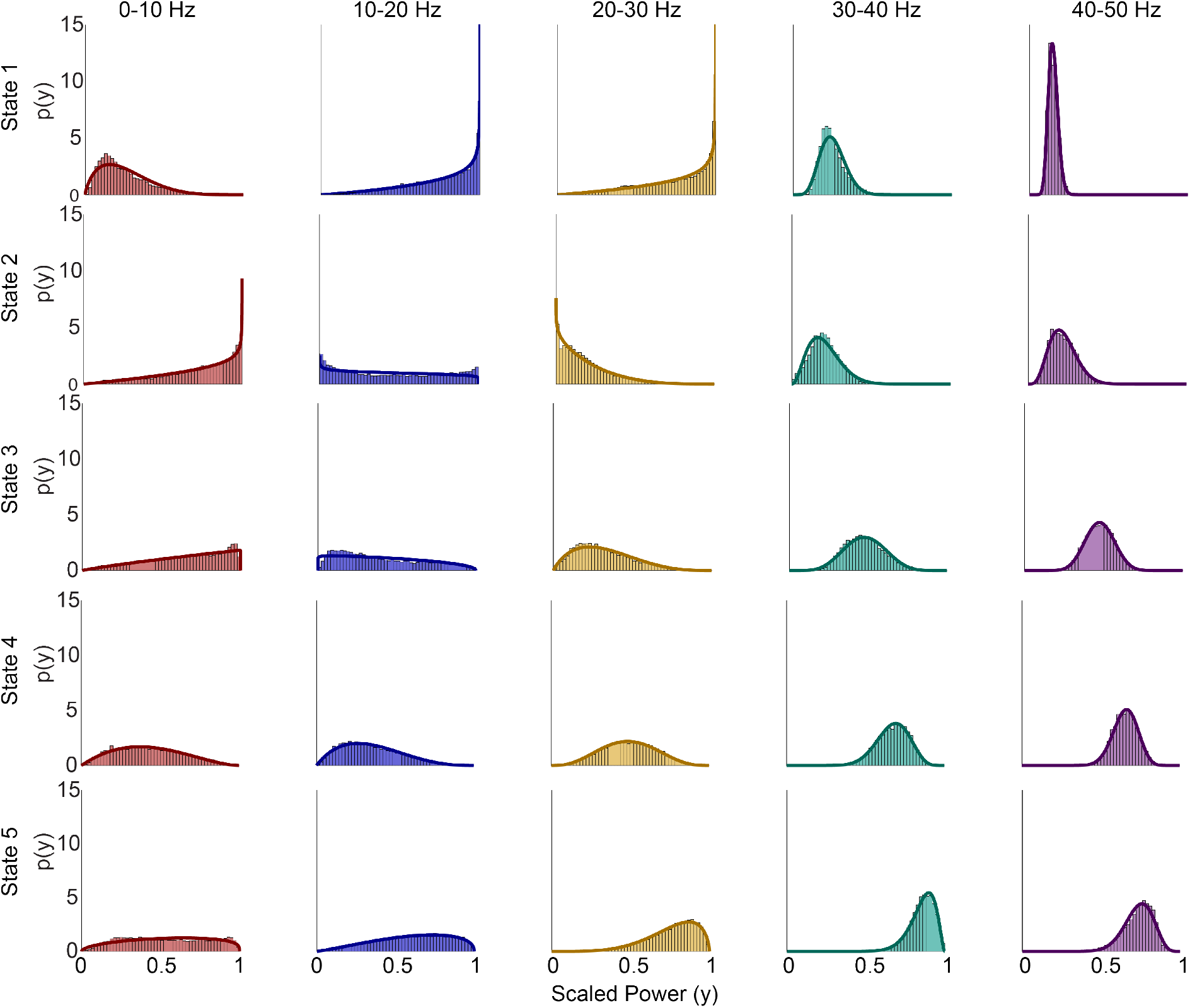
Illustration of the state-specific and frequency-band specific beta pdfs corresponding to a 5-state beta-HMM that is estimated from *L* = 4 sessions of LFP recording from NHP MJ. For each state (viewed row-wise) and a frequency band (viewed column-wise), the corresponding subplot presents (1) the empirical pdf plotted as a histogram based on the observations that correspond to the optimal segmentation across the L=4 sessions, and (2) the continuous beta pdf with parameters estimated by the EM algorithm. Each color distinguishes a frequency band.

**Figure 12:**
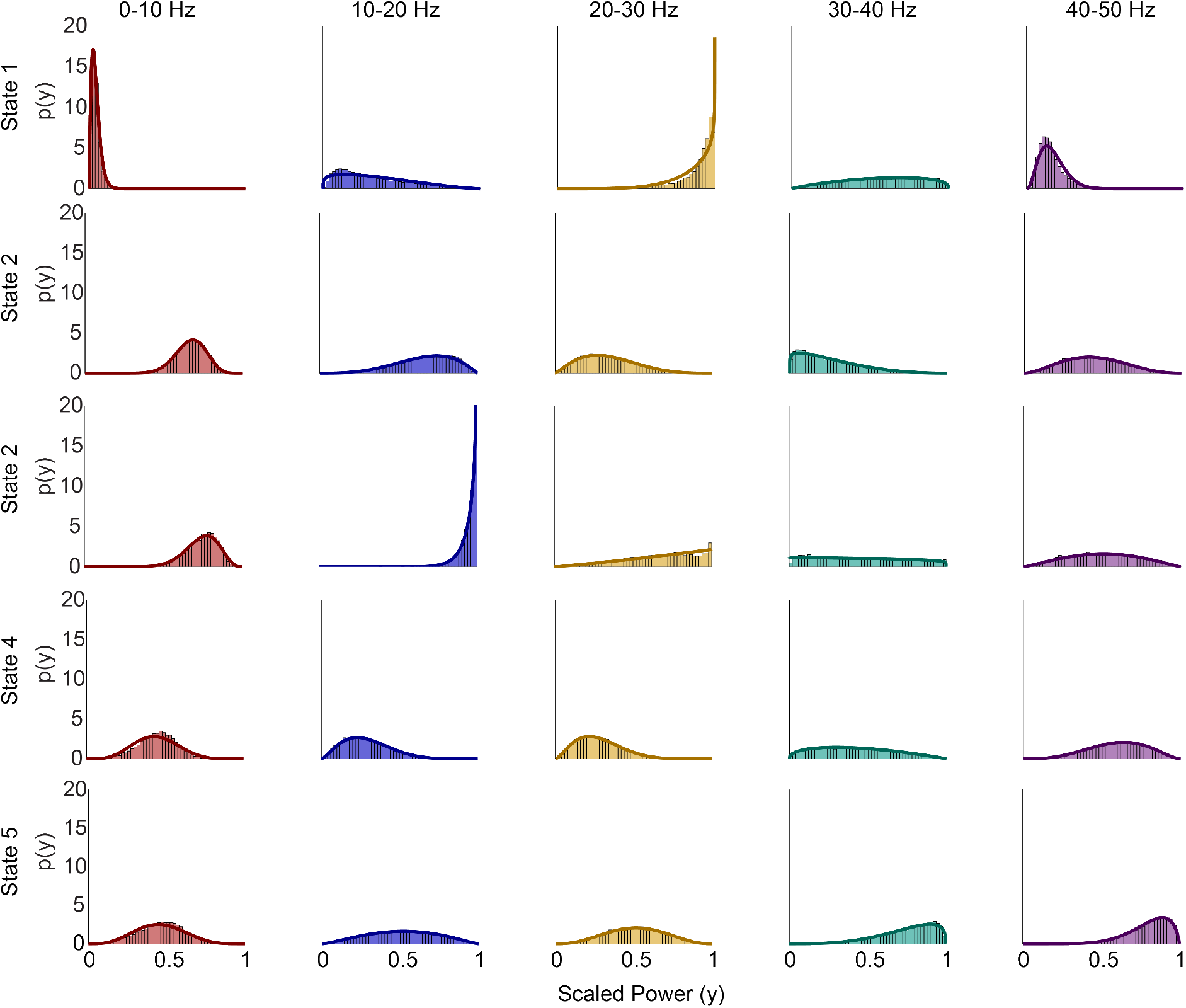
Illustration of the state-specific and frequency-band specific beta pdfs corresponding to a 5-state beta-HMM that is estimated from *L* = 5 sessions of LFP recording from NHP LM. For each state (viewed row-wise) and a frequency band (viewed column-wise), the corresponding subplot presents (1) the empirical pdf plotted as a histogram based on the observations that correspond to the optimal segmentation across the L=5 sessions, and (2) the continuous beta pdf with parameters estimated by the EM algorithm. Each color distinguishes a frequency band.

We use the subject-specific HMMs to further analyze the spectral content and duration statistics across states, both within a subject as well as between the two subjects. We use the ML estimated beta distributions to perform pairwise comparisons of the scaled power between any two states from either NHP (Fig. 5(A-D)). By investigating *Pr*(Δ_*hjk*_ ≤ 0; *a*_*hj*_, *b*_*hj*_, *a*_*hk*_, *b*_*hk*_) (per Sec. 2.5), we were able to determine the probability that HMM state *j* had lower scaled power than HMM state *k* in the *h*-th frequency band. For this section, we simplify the notation by referring to *Pr*(Δ_*hjk*_ ≤ 0; *a*_*hj*_, *b*_*hj*_, *a*_*hk*_, *b*_*hk*_) as *Pr*(Δ_*hjk*_ ≤ 0), and summarize the key findings^5^. In both NHPs, we found that the scaled powers in the 0 − 10 Hz and 40 − 50 Hz bands during the pre-ketamine state (HMM state 1) were lower relative to every other state (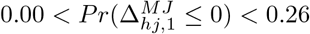 and 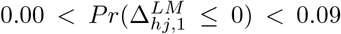 for *h* ∈ {1, 5} and *j* ∈ [2, *K*]). This provides further evidence that the oscillatory dynamics in these two frequency bands were most significantly altered by ketamine. NHP MJ had higher scaled power in the 30-40 and 40-50 Hz ranges in states 4 and 5 compared to states 2 and 3 (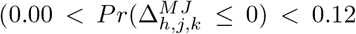 for *h* ∈ [4, 5], *j* ∈ [4, 5], and *k* ∈ [2, 3]). This is consistent with the observation that the gamma bursts (represented by states 4 and 5) in NHP MJ occurred broadly between 30-50 Hz. NHP LM also had higher scaled power in the 40-50 Hz range in states 4 and 5 compared to states 2 and 3 (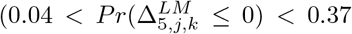 for *j* ∈ [4, 5], and *k* ∈ [2, 3]). However, state 4 in NHP LM did not have higher 30-40 Hz scaled power than state 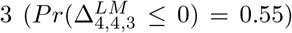. This is consistent with the observation that the gamma bursts (represented by states 4 and 5) occurred primarily between 40-50 Hz in NHP LM. In both NHPs, state 2 had higher scaled power in the 0-10 Hz range compared to states 4 and 5 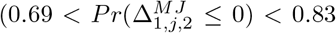 and 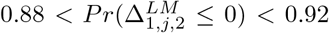 for *j* ∈ [4, 5]), and lower power in the 30-40 and 40-50 Hz ranges compared to states 3 to 5 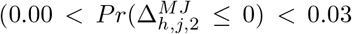 and 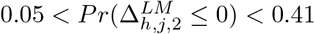 for *h* ∈ [4, 5] and *j* ∈ [3, 5]).

We further leveraged the subject-specific beta-HMMs to compare beta distributions between any HMM state in NHP MJ and any HMM state in NHP LM (see Sec. 2.5, Fig. 5(C)). Fig. 5(D) shows the diagonal of each of the matrices in Fig. 5(C), and summarizes how each state in NHP MJ compares to the corresponding state in NHP LM. In the 40 − 50 Hz range, we found that HMM states 3, 4, and 5 in NHP MJ were most similar to the HMM states 3, 4, and 5 in NHP LM, respectively 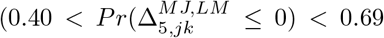 for *j* ∈ [3, 5] and *k* = *j*). State 2 in NHP MJ was more similar to state 1 in NHP LM in the 40-50 Hz band 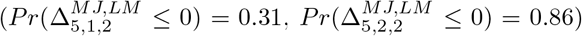, while state 2 in NHP LM was more similar to state 3 in NHP MJ 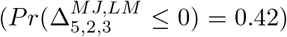. This indicates that following a ketamine bolus, both NHPs have similar activity in the 40-50 Hz band, but that the suppression of gamma activity in state 2 in NHP MJ is more prominent than in NHP LM.

We found significant differences in the neurophysiological state dynamics between the two primates. From the transition matrices, it is apparent that the mean duration in each state in NHP MJ is greater than that of NHP LM, indicating faster state-switching dynamics in the latter subject. For NHP MJ, the transition matrix revealed that the neurophysiological states switch between the slow oscillation (HMM state 2) and the gamma burst states (HMM states 4 and 5), moving via an intermediate transition state (HMM state 3). In contrast, for NHP LM, the slow oscillation state is more likely to transition directly to the gamma burst states and vice versa. The mean duration of the gamma burst state, 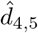 (i.e. consecutive time-windows spent in states 4 or 5), was 2.5*s*([1.9, 3.4]*s*) in NHP MJ, and 1.2*s*([0.9, 1.5]*s*) in NHP LM. The mean interval between consecutive occurrences of gamma burst states, 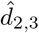, was 2.2*s*([1.5, 3.3]) in NHP MJ, and 1.1*s*([0.8, 1.3]*s*) in NHP LM. The mean duration of the slow oscillation state, 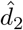, was 1.6*s*([1.1, 2.5]*s*) in NHP MJ, and 0.7*s*([0.6, 0.9]*s*) in NHP LM. The mean interval between consecutive occurrences of slow oscillation states, 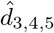, was 6.6*s*([4.0, 11.9]*s*) in NHP MJ and 1.7*s*([1.3, 2.2]*s*) in NHP LM.

### 4.4 Human-specific beta-HMM estimated using observations from multiple patient subjects (*L >* 1 case)

We investigated if the beta-HMM analysis framework could be applied to infer the common ketamine-induced neurophysiological state dynamics observed in the scalp EEG of multiple OR patients. Therefore, we fit a single beta-HMM to independently collected EEG data from *L* = 9 human patients and generated a quantitative description of ketamine-induced neurophysiological dynamics for a typical patient (Fig. 6). For this analysis, we chose a model order of *K* = 6 states, which allowed for classification of broadband high power, typical of noise artifacts in the OR, as an additional HMM state.

As with NHPs, we found that the beta distributions in each frequency band were unique across all states (KS-distance *>* 0.07 for all pairwise comparisons; 75 comparisons = 15 pairs of states per frequency band × 5 frequency bands) (Fig. 6(J); also see Figs. 13 in the Supporting Figures section). We found that HMM states 4 and 5 had high scaled power in the 30-40 Hz and 40-50 Hz frequency ranges (*Pr*(*Y*_4_ *>* 0.5|*Z* = 4) = 0.79, *Pr*(*Y*_4_ *>* 0.5|*Z* = 5) = 0.99, *Pr*(*Y*_5_ *>* 0.5|*Z* = 4) = 0.66, *Pr*(*Y*_5_ *>* 0.5|*Z* = 5) = 0.91). We again considered states 4 and 5 to represent the neurophysiological gamma burst state. There were also some distinct neurophysiological differences between the human EEG and the NHP LFP. First, there was a strong slow-oscillation throughout the human EEG recordings, which resulted in more overlap between the pre-ketamine HMM states and the post-ketamine HMM states in the frontal electrodes. For this reason, identifying a state dominated by slow activity was less clear in the human beta-HMM model compared to the NHP beta-HMM models. However, states 1 and 2 had low power in the 30-40 Hz and 40-50 Hz frequency ranges (*Pr*(*Y*_4_ *>* 0.5|*Z* = 1) = 0.00, *Pr*(*Y*_4_ *>* 0.5|*Z* = 2) = 0.04, *Pr*(*Y*_5_ *>* 0.5|*Z* = 1) = 0.00, *Pr*(*Y*_5_ *>* 0.5|*Z* = 2) = 0.21), which is consistent with our definition of the neurophysiological slow oscillation state, characterized by strong slow activity and low high frequency activity. Second, the gamma bursts in humans were characterized by a more broadband increase in power – state 5 also had high power in the 10-30 Hz frequency ranges (*Pr*(*Y*_2_ *>* 0.5|*Z* = 5) = 0.76, *Pr*(*Y*_3_ *>* 0.5|*Z* = 5) = 0.98). Finally, soon after the time of the ketamine bolus, in patients A, C, E, F, G, and H, there was broadband high power that, in the context of OR cases, is difficult to distinguish from noise. This activity is classified with the 6-th state, which also corresponds to periods of obvious noise artifacts (e.g. at t = 0 min in patient D).

**Figure 13:**
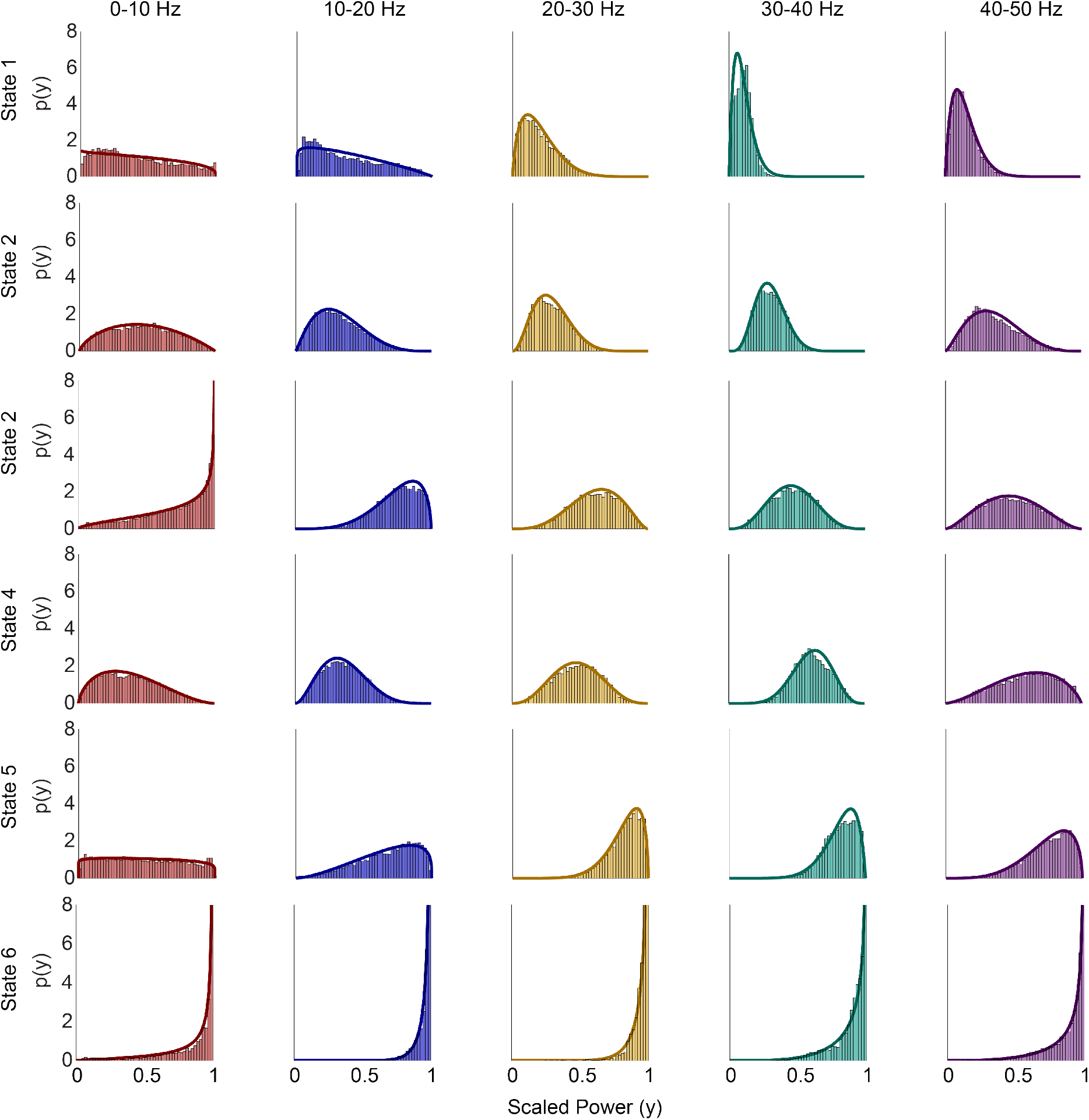
Illustration of the state-specific and frequency-band specific beta pdfs corresponding to a 6-state beta-HMM that is estimated from *L* = 9 sessions of EEG recording from 9 human OR patients. For each state (viewed row-wise) and a frequency band (viewed column-wise), the corresponding subplot presents (1) the empirical pdf plotted as a histogram based on the observations that correspond to the optimal segmentation across the L=9 sessions, and (2) the continuous beta pdf with parameters estimated by the EM algorithm. Each color distinguishes a frequency band.

Further quantitative analysis (Fig. 7) of human EEG beta-HMM parameters revealed high scaled power in the 30-50 Hz range for the states characterized by gamma burst activity 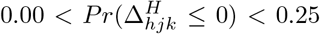 for *h* ∈ [4, 5], *j* ∈ [4, 5] and *k* ∈ [1, 3]). As previously noted, there was strong slow oscillation activity throughout the human recordings, and so the changes in 0-10 Hz for states 1-5 were more subtle than in the NHPs. States 1 and 2 had the lowest power between 30-50 Hz 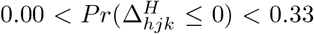 for *h* ∈ [4, 5], *j* ∈ [3, 5], and *k* ∈ [1, 2]). Thus, HMM states 1 and 2 best represent the neurophysiological slow oscillation state, characterized by strong slow activity and weak high frequency activity. Finally, through this analysis, it is again clear that state 6 was characterized by broadband high power 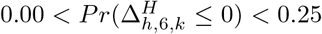 for *h* ∈ [1, 5] and *k* ∈ [1, 5]).

Quantitative analysis of the beta-HMM dynamics revealed that the human EEG also tends to transition cyclically through the HMM states induced by ketamine – the most probable transition sequence, starting from state 1, is 1 → 2 → 4 → 5 → 3 → 2. All states have a low probability of transitioning to state 6 (*A*_*j*,6_ *<* 0.008 for *j* ∈ [1, 5]). Furthermore, the mean duration of the gamma burst state, 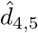, was 2.7*s*([1.9, 3.8]*s*). The mean interval between consecutive occurrences of the gamma burst state, 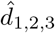, was 3.8*s*([2.5, 5.8]*s*). The mean duration of the slow oscillation state 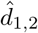 was 2.8*s*([1.9, 4.3]*s*). The mean interval between consecutive occurrences of slow oscillation state 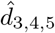 was 3.6*s*([2.5, 5.4]*s*).

## 5 Discussion

Our beta-HMM analysis framework provided parsimonious summaries of ketamine-induced slow-gamma alternating dynamics for NHP LFP and human EEG recordings following high-dose ketamine administration. The input to the analysis framework was one or more sequences of instantaneous power in multiple frequency bands. The power values were averaged across frequencies within each discrete band and scaled between 0 and 1. The scaling enabled identification of latent HMM states each with a distinct probability distribution on power values lying between a very low (corresponding to a 0) and a very high value (corresponding to a 1) in each of the chosen frequency bands, irrespective of the absolute value of spectral estimates. A key assumption incorporated in the HMM is that at any instant in time, the probability distribution from which the observation is sampled depends only on the discrete value of an underlying latent state process at the same instant. This state process itself is assumed to evolve according to a first-order Markovian transition dynamics. The key outputs of our analysis comprised the model parameters estimated using an EM algorithm and a corresponding state trajectory estimated using the Viterbi algorithm. The state trajectory provided an objective and efficient segmentation of LFP and EEG data. The estimated state-specific and frequency band-specific beta pdf’s characterized the spectral representation of each HMM state, whereas the transition matrix characterized the underlying dynamics. Using the estimated observation pdf’s, we objectively defined neurophysiological states (e.g. gamma burst state, slow oscillation state) in terms of one or more HMM states. Furthermore, by using the estimated state transition matrices, we calculated mean duration and mean interval statistics (as defined in Sec. 2.5) corresponding to each of these neurophysiological states.

Our analyses revealed an alternating pattern of states characterized primarily by gamma burst and slow oscillation activity, as well as intermediate state in between. The mean duration of the gamma burst state was 2.5s ([1.9, 3.4]s) and 1.2s ([0.9, 1.5]s) for the two NHPs, and 2.7s ([1.9, 3.8]s) for the nine human subjects. The mean duration of the slow oscillation state was 1.6s ([1.1, 2.5]s) and 0.7s ([0.6, 0.9]s) for the two NHPs, and 2.8s ([1.9, 4.3]s) for the nine human subjects. Thus, our objective beta-HMM analysis framework enabled us to precisely characterize the dynamics of ketamine-induced gamma burst and slow oscillation states, and to compare the time-scales of these neurophysiological states between subjects and between species.

Our work advances the development and application of LFP or EEG data analysis tools. The beta-HMM analysis framework falls under the broad category of research aimed at developing time-frequency state space models to analyze neural dynamics [41, 50, 64, 65, 66, 67], and applying them in principled interpretation of neural time-series data for unconsciousness research [12, 64]. A few key distinguishing features of the beta-HMM analysis framework are as follows. The assumption of state-specific and frequency-band specific beta distributions as marginal distributions of the instantaneous observation vectors allows for a flexible framework to characterize highly variable spectral profiles. This assumption, along with the assumption of Markovian state transitions, allowed us to directly characterize the discontinuous switching among multiple spectral profiles, as well as identify segments with similar spectral properties (See figures 3 C, D; 4 F, M; and 6 K). Furthermore, our beta-HMM analysis framework accounts for the disjoint nature of data from separate sessions without requiring any concatenation of sessions or consequently any arbitrary choice of concatenation order. Potential extensions of this work on the statistical methods development front may include modeling the transition matrix as time-varying, which would account for longer time-scale variations (e.g. varying drug-levels). Furthermore, generalizing the current single-channel LFP or EEG data-based analysis framework to incorporate multi-channel LFP or EEG data would allow for neurophysiological characterization of ketamine-induced altered arousal states based on spatio-temporal statistical relationships in the observed data. Going beyond the context of ketamine-induced altered arousal states, the analysis framework itself can be relevant in general use-cases where LFP or EEG data demonstrate discontinuous, repeating transitions among time-limited, band-limited spectral signatures, such as those observed during sleep [41, 68] or burst suppression in general anesthesia [69, 70]. In such applications, it will also be straight forward to modify the definitions of frequency bands in our work to incorporate frequency bands conventionally adopted in neuroscience studies [11, 71]^6^.

Our beta-HMM research can inform future neuroscience inquiries. While there are several hypotheses for the generation of gamma activity under ketamine [19], there is not yet a clear understanding of how the bursting activity is generated. Preliminary work indicates that there may be a cycle of cortical inhibition and dis-inhibition due to activity-dependent ketamine NMDAR inhibition of cortical interneurons and pyramidal neurons [20]. Our analyses of the NHP datasets using the beta-HMM led to the identification of neurophysiological states that were primarily associated with the period following administration of a ketamine bolus. The summary statistics from each NHP inform *which* frequency bands show significant activity, and *how* this activity varies over time. The duration and interval statistics of the alternating gamma burst state, and the slow oscillation states that we estimated for NHP LFP can serve as quantitative constraints in the design of rhythm-generating neuronal microcircuit models that would mimic the neurophysiological dynamics caused by ketamine, similar to ones previously done for other anesthetics [21, 22, 24, 72, 73]. Furthermore, we identified a previously unreported transition state between the alternating gamma burst and slow oscillation states, and a mechanistic characterization of this state would complement ongoing statistical analyses of neurophysiological data.

Our beta-HMM research can also inform future clinical inquiries. Ketamine is known to produce distinct oscillatory signatures in the electroencephalogram (EEG) of healthy volunteers and patients [10, 11]. These oscillatory signatures are starkly different from those produced during propofol-induced unconsciousness [71, 74]. These differences in oscillatory dynamics, particularly the presence of alternating slow oscillation and gamma burst states under ketamine, indicate that a spectral marker of propofol-induced unconsciousness is not a reliable marker for tracking ketamine-induced altered arousal states. However, the existence of consistent neurophysiological states observed in EEG following ketamine bolus administration in multiple patients indicate promise in precise monitoring of these states to track ketamine-induced altered states of arousal. Towards this goal, as demonstrated in this work, the beta-HMM analysis framework can provide precise, objective characterization of the neurophysiological dynamics associated with ketamine-induced altered states of arousal. Furthermore, in addition to its use for general anesthesia and analgesia, ketamine is also used in treatment of depression [75, 76] and in pharmacologic models for schizophrenia [18, 77, 78]. Thus the clinical utility and mechanistic insights gained by detailed quantification of the neural activity corresponding to ketamine-induced altered states of arousal can potentially aid future research and clinical applications.

There are two primary limitations in both the data set that we analysed here and in our assumptions. The first limitation is a need for prospective studies in humans and animals with carefully recorded simultaneous neural, behavioral response and physiological activities. Future experimental studies with larger cohorts can address the current study’s limitation due to low number of subjects in both the NHP LFP and human EEG analyses. The consistency of the alternating gamma-slow spectral activity from 9 patients and 2 NHPs during ketamine-induced altered arousal states indicate that transition dynamics between the gamma burst and slow-delta neurophysiological states can be a candidate biomarker of ketamine-induced unconsciousness. However, to reliably establish these EEG or LFP correlates of unconsciousness, experimental studies in healthy volunteers and animal subjects with simultaneous monitoring of behavioral response, physiological parameters and neural activity will be essential. The second limitation is a need for extension to realtime tracking of neurophysiological states during ketamine-induced general anesthesia. While our current approach utilizes the entire data set to calculate appropriate scaling factors for transforming spectral power to real numbers lying between 0 and 1, and thus (in the present formulation) cannot be executed in real time. A larger study may allow for the development of a generalized scaling approach. This would facilitate objective identification of ketamine neurophysiological states in real-time. While a common limitation of EEG analysis in real-time OR settings is the presence of motion and other noise artifacts in recordings, the beta-HMM framework is well poised to handle these artifacts, as observations that fall far outside the distribution of the signal can be assigned to a distinct HMM state.

In conclusion, we have developed a detailed analysis framework for ketamine-induced neurophysiological phenomena. The generalizability of the beta-HMM framework indicates utility beyond what has been reported here. Future work will investigate how the spectral dynamics revealed by our analysis contribute to ketamine-induced altered states of arousal.

## Data Availability

Data and code will be made publicly available when the paper is published.

## 6 Acknowledgement

The authors thank Prof. Polina Anikeeva for her feedback throughout the manuscript preparation and Dr. Alexis Garcia for suggesting helpful references.

## 8 Author Contributions

**Conceptualization**: S. Chakravarty, I. C. Garwood, E. N. Brown

**Formal Analysis**: I. C. Garwood, S. Chakravarty, E. N. Brown

**Funding Acquisition**: E. N. Brown

**Investigation**: I. C. Garwood, S. Chakravarty, J. Donoghue, O. Akeju

**Methodology**: I. C. Garwood, S. Chakravarty

**Software**: I. C. Garwood, S. Chakravarty

**Resources**: E. K. Miller, O. Akeju, E. N. Brown

**Data Curation**: I. C. Garwood, J. Donoghue, S. Chamadia, O. Akeju, P. Kahali

**Supervision**: E. N. Brown

**Visualization**: I. C. Garwood

**Writing-Original Draft**: I. C. Garwood, S. Chakravarty

**Writing-review & editing**: I. C. Garwood, S. Chakravarty, E. N. Brown, E. K. Miller, S. Chamadia, O. Akeju, J. Donoghue, P. Kahali

## A M-step of EM

We calculate Θ^(*n*)^ by solving the constrained maximization of the 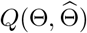 (Eq. (8)),

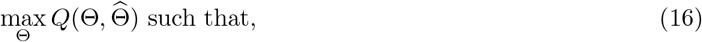

Equality constraints:

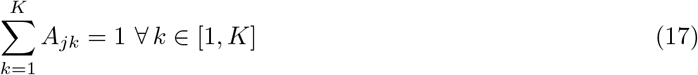

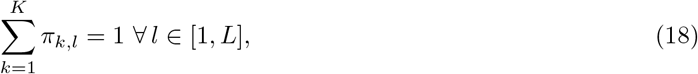

Inequality constraints:

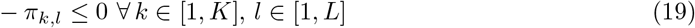

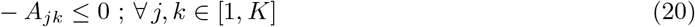

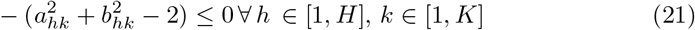

Due to the independent additive contributions to the objective function due to each group of decision variables, *π*_*k,l*_, {***A*** and 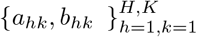, we seek to increase 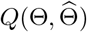 by maximizing these individual contributions independently. To implement this we solve the following optimization problems,

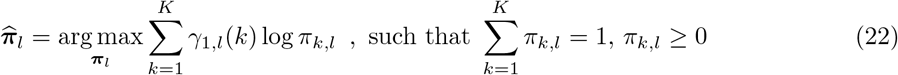

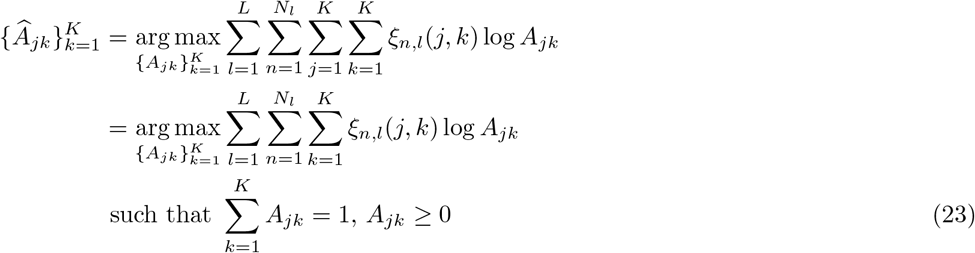

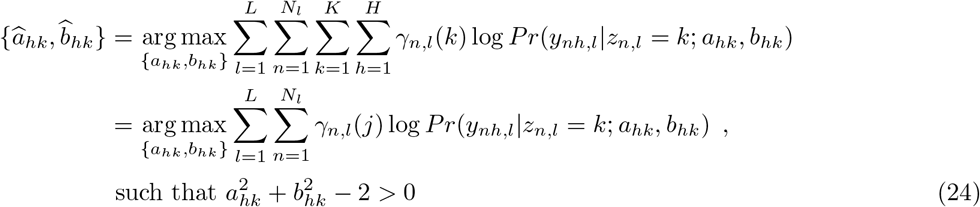

Note that in Eqs. (22), (23) and (24) we are solving for 1, *K* and *KH* independent optimization problems. The solutions to Eq. (22) and the (23) has the following closed-form expressions,

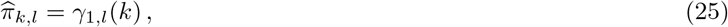

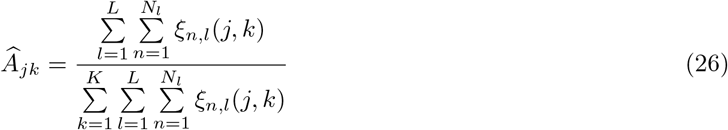

To solve the constrained optimization problem in Eq. (24), we use the Lagrange multiplier method [79]. Alternatively, we can directly employ numerical tools for constrained optimization such as the `fmincon` function in Matlab.

## B Tutorial on analyzing HMM state-specific spectra in terms of corresponding estimated beta pdf’s

Here we present a tutorial-styled discussion on how to use HMM state-specific frequency band-specific beta pdf’s (estimated using EM algorithm described in Sec. 2.4) to interpret the corresponding HMM state-specific spectral profiles. To demonstrate the utility of the state-specific beta distributions, we use segmented data from the 2 HMM states, states 2 and 5, identified in the single session analysis of NHP LFP data (Fig. 3). To elucidate the correspondence between the spectral profile (Fig. 8(A, B) and Fig. 8(E,F)) and the respective empirical pdfs of the scaled powers (Fig. 8(C) and Fig. 8(G)), we recall the intuition provided in Sec 2.2 with regard to the scaling equation (Eq. (2)). Recall that the scaling factor for each frequency band is calculated from the quartiles of the spectrogram across the entire session. As a result, if the distribution of spectral power corresponding to a given state and frequency band has a similar variance to that of the entire session, its scaled representation will have high variance. This will correspond to a relatively flat beta distribution. In contrast, if the spectral power from a given state and frequency band tends to be greater than the median of the corresponding frequency band, the scaled observations will be skewed towards 1. Particularly, if the observations corresponding to a given HMM state are mostly between the third and fourth quartile, the pdf of the beta distribution for that state will have high probability mass between 0.75 and 1. Note that corresponding statements can be made for states with relatively lower spectral power in a given frequency band (i.e., the beta distributions would be skewed towards 0). We use this intuition to quantitatively characterize the spectral power in a given frequency band for states 2 and 5.

As seen in Fig. 8(A,B), there is prominent power in the 0-10 Hz band for state 2. These power values are higher than the median value across the entire session, as indicated by high probability mass near 1 in Fig. 8(C). This right shift of the probability mass towards 1 in the scaled power pdf’s is also represented by a high complementary cdf (ccdf)^7^ at 0.5 (Fig. 8(D)) with *Pr*(*Y*_1_ > 0.5|*Z* = 2) = 0.82. In state 2, there is not much activity in the 10-20 Hz band (Fig. 8(B)). This is similar to the activity across the majority of the session, and so the distribution of the scaled power has high variance (Fig. 8(C); our analysis of all the states in Fig 3 also show that 10-20 Hz power is not a discriminating feature across the states). In state 2, there is not much activity in the 20-30Hz, 30-40 Hz, and 40-50 Hz bands (Fig. 8(B)), and the power values are mostly lower than the median (as indicated by high probability mass near 0 in Fig. 8(C)). The left shift of the probability mass towards 0 in the scaled power is also represented by the low ccdf values at 0.5 (Fig. 8(D)) with *Pr*(*Y*_3_ > 0.5|*Z* = 2) = 0.002, *Pr*(*Y*_4_ > 0.5|*Z* = 2) = 0.002, and *Pr*(*Y*_5_ > 0.5|*Z* = 2) = 0.004.

Now using the intuitions built from our discussion of state 2, let us try to interpret the spectral representation of state 5. In the 0-10 Hz band there is prominent activity in state 5 (Fig. 8(E,F)), but the high variance of the distribution of scaled power (Fig. 8(G)) indicates that the variance of the unscaled power from state 2 is similar to the variance across the entire session. In 10-20 Hz band there is not much activity in state 5 (Fig. 8(F)), but the power values tend to be higher than the median value of the entire session (as indicated by the large spread but a mode > 0.5 in Fig. 8(G)). In each of the 20-30Hz, 30-40Hz, 40-50Hz bands there is prominent activity (Fig. 8(E,F)), and the large probability mass near 1 in the scaled power pdf’s (Fig. 8(G)) indicates that these activities are higher than the respective median values. This right shift is clearly captured by the ccdf values at 0.5 – *Pr*(*Y*_3_ > 0.5|*Z* = 5) = 0.98, *Pr*(*Y*_4_ > 0.5|*Z* = 5) = 1.00, and *Pr*(*Y*_5_ > 0.5|*Z* = 5) = 0.97.

We can further use these beta pdfs to quantitatively compare the power values in a given frequency band between states 2 and 5. Using the Eq. (9), we define a Δ_*h*,5,2_ = *X*_*h*,5_–*X*_*h*,2_, such that *X*_*h*,5_ ∼ *Beta*(*a*_*h*,5_, *b*_*h*,5_) and *X*_*h*,2_ ∼ *Beta*(*a*_*h*,2_, *b*_*h*,2_). Fig. 8(I) and Fig. 8(J) represent the pdf and cdf for the Δ_*h*,5,2_ where *h* ∈ [1, 5]. For bands 0-10 Hz and 10-20 Hz, most of the probability mass on Δ_*h*,5,2_ lies around 0 and the corresponding CDF values at 0 (which correspond to *Pr*(Δ_*h*,5,2_ ≤ 0)) are 0.70 and 0.34, respectively. This indicates that while state 2 tends to have higher power in the 0-10 Hz band and state 5 tends to have higher power in the 10-20 Hz band, the activity in these bands is most likely not a significant discriminating factor between the two states. However, for the bands 20-30Hz, 30-40Hz, 40-50Hz, most of the probability mass of the respective Δ’s occurs to the right of 0, which results in CDF values at 0 of 0.01, 1.9 × 10^−5^ and 1.1 × 10^−4^, respectively. This indicates that the power in these three frequency bands is higher in state 5 compared to state 2, and the activity in these bands is likely to be significant discriminating factor between the two states. Thus we can generate a quantitative description to support our qualitative assessment that in high frequency bands, state 5 has higher power relative to state 2.

## C Supporting figures and table

## Footnotes

1 We use bold math symbols to indicate vector-valued quantities.

2 A general approach to calculate probability distributions for difference random variables can be found in the work by Cook and Nadarajah[60]

3 See Sec B for a tutorial-styled explanation of how the estimated beta pdf’s are used to analyze the spectral properties of the HMM states

4 See Sec B for a tutorial-styled explanation of how the estimated beta pdf’s are used to analyze the spectral properties of the HMM states

5 See Sec B for a tutorial-styled explanation of how the estimated beta pdf’s are used to analyze the spectral properties of the HMM states

6 Reiterating, the motivation for our choice of frequency bands in this work was to maintain consistency and objectivity in our analyses across the multiple NHP LFP and human EEG datasets.

7 ccdf at a given sample point equals 1 minus the value of the cdf at the same sample point

